# Data-driven consideration of genetic disorders for global genomic newborn screening programs

**DOI:** 10.1101/2024.03.24.24304797

**Authors:** Thomas Minten, Sarah Bick, Sophia Adelson, Nils Gehlenborg, Laura M. Amendola, François Boemer, Alison J. Coffey, Nicolas Encina, Alessandra Ferlini, Janbernd Kirschner, Bianca E. Russell, Laurent Servais, Kristen L. Sund, Ryan J. Taft, Petros Tsipouras, Hana Zouk, ICoNS Gene List Contributors, David Bick, the International Consortium on Newborn Sequencing (ICoNS), Robert C. Green, Nina B. Gold

**Affiliations:** KU Leuven; Boston Children’s Hospital; Massachusetts General Hospital; Harvard Medical School; Brigham and Women’s Hospital; Stanford School of Medicine; Harvard Medical School, Department of Biomedical Informatics; National Institutes of Health, National Institute of Allergy and Infectious Disease; University of Liege, CHU Liege; Illumina Inc; ICoNS; Ariadne Labs; Harvard T.H. Chan School of Public Health; University of Ferrara, Department of Medical Sciences, Department of Medical Sciences, Unit of Medical Genetics; University Medical Center Freiburg, Department of Neuropediatrics and Muscle Disorders; University of California, Los Angeles, David Geffen School of Medicine, Department of Human Genetics, Division of Clinical Genetics; University of Oxford; University of Liege; Nurture Genomics; FirstSteps-BNSI; Massachusetts General Hospital, Department of Pathology, Laboratory for Molecular Medicine; Harvard Medical School, Department of Pathology; Broad Institute; Genomics England; Mass General Brigham; Massachusetts General Hospital, Department of Pediatrics; Harvard Medical School, Department of Pediatrics

## Abstract

**Purpose:** Over 30 international studies are exploring newborn sequencing (NBSeq) to expand the range of genetic disorders included in newborn screening. Substantial variability in gene selection across programs exists, highlighting the need for a systematic approach to prioritize genes.

**Methods:** We assembled a dataset comprising 25 characteristics about each of the 4,390 genes included in 27 NBSeq programs. We used regression analysis to identify several predictors of inclusion, and developed a machine learning model to rank genes for public health consideration.

**Results:** Among 27 NBSeq programs, the number of genes analyzed ranged from 134 to 4,299, with only 74 (1.7%) genes included by over 80% of programs. The most significant associations with gene inclusion across programs were presence on the US Recommended Uniform Screening Panel (inclusion increase of 74.7%, CI: 71.0%-78.4%), robust evidence on the natural history (29.5%, CI: 24.6%-34.4%) and treatment efficacy (17.0%, CI: 12.3%- 21.7%) of the associated genetic disease. A boosted trees machine learning model using 13 predictors achieved high accuracy in predicting gene inclusion across programs (AUC = 0.915, R² = 84%).

**Conclusion:** The machine learning model developed here provides a ranked list of genes that can adapt to emerging evidence and regional needs, enabling more consistent and informed gene selection in NBSeq initiatives.

## Introduction

A decade ago, the BabySeq Project piloted newborn and childhood sequencing (NBSeq), a process designed to detect risk for a wide range of genetic disorders in apparently healthy infants.^1–9^ Per the data repository Rx-Genes, over 700 genetic disorders now have targeted treatments or consensus guidelines for long-term management, which has further fueled enthusiasm for NBSeq.^10,11^ Stakeholders, including diverse groups of parents,^12–14^ rare disease specialists,^11^ primary care physicians,^15^ genetic counselors,^11,16,17^ and the public^18,19^ now support the implementation of genomic newborn screening for at least some disorders. At least 30 international research programs and companies are actively exploring this screening approach,^20–22^ most of which are exchanging best practices through the International Consortium on Newborn Sequencing (ICoNS).^23^

Historically, the criteria established by Wilson and Jungner^24^ have provided a framework for selecting the disorders to include in public newborn screening programs. These criteria prioritize the inclusion of childhood-onset disorders that are treatable if diagnosed in their earliest stages and require immediate intervention to prevent irreversible damage. Given the hundreds of treatable disorders that could be candidates for NBSeq and the complexity of genomic data, however, selection of the appropriate genes and disorders for NBSeq is a recurring challenge.^21,25,26^ Prior studies have identified discrepancies across the genes being analyzed by a limited number of commercial NBSeq programs^27^ and research studies,^28,29^ but little is known about the values and variables that underlie these differences. Understanding which genes have high concordance across programs may guide emerging NBSeq research programs as they select which genes and variants to report to participants. Furthermore, the characteristics of these genes and their associated disorders can be used more empirically to prioritize candidate genes for public health programs.

To understand the variability among newborn sequencing programs, we compared the genes currently selected for analysis by 27 research studies and commercial NBSeq programs. For each gene that was included in any NBSeq program, we assembled a dataset of 25 associated characteristics. We then used a multivariate regression analysis to identify which of these characteristics were associated with inclusion across programs. Finally, we used a boosted trees model to generate a ranked list of genes, offering a data-driven approach to the prioritization of genetic disorders for population-wide NBSeq.

## Materials and methods

### Study design

This cross-sectional study involved four stages: (1) identification of gene lists across international NBSeq research studies and commercial programs, (2) compilation of a dataset of characteristics for each gene included in any program, (3) statistical comparison of gene lists across programs, and (4) development of a machine learning model to predict gene inclusion across NBSeq programs based on gene characteristics.

### Identification of lists of genes from research studies and commercial programs

A total of 35 NBSeq programs were identified through membership in the International Consortium on Newborn Sequencing (ICoNS) and an online search (Supplementary Table S1, Figure 1).^5,30–47^ We obtained gene lists from 27 NBseq programs, including 20 research studies: BabyDetect,^30,31^ BabyScreen+,^28^ BabySeq,^2^ BeginNGS,^32,33^ Chen et al. 2023,^34^ Early Check,^35,48^ FirstSteps, the Generation study, gnSTAR,^36,38^ GUARDIAN study,^42,49,50^ Jian et al. 2022,^37^ Lee et al. 2019,^43^ Luo et al. 2020,^44^ NeoExome,^47^ NeoSeq,^40^ NESTS,^41^ NewbornsInSA, Progetto Genoma Puglia, Screen4Care,^45^ and Wang et al. 2023.^39^ Seven lists of genes from commercial firms that offer genomic newborn screening were included: FORESITE 360 (Fore Genomics), Fulgent (Newborn Genetic Analysis), Igenomix (Igenomix newborn screening), Mendelics (Teste da Bochechinha), Nurture Genomics, PerkinElmer Revvity (Genetic InsightPanel),^46^ and Sema4 (Natalis).^30^ Nine studies reported screening outcomes and we also summarized the positive screen rates and positive predictive values for these studies (Table 1, Supplementary Methods).

**Figure 1.**
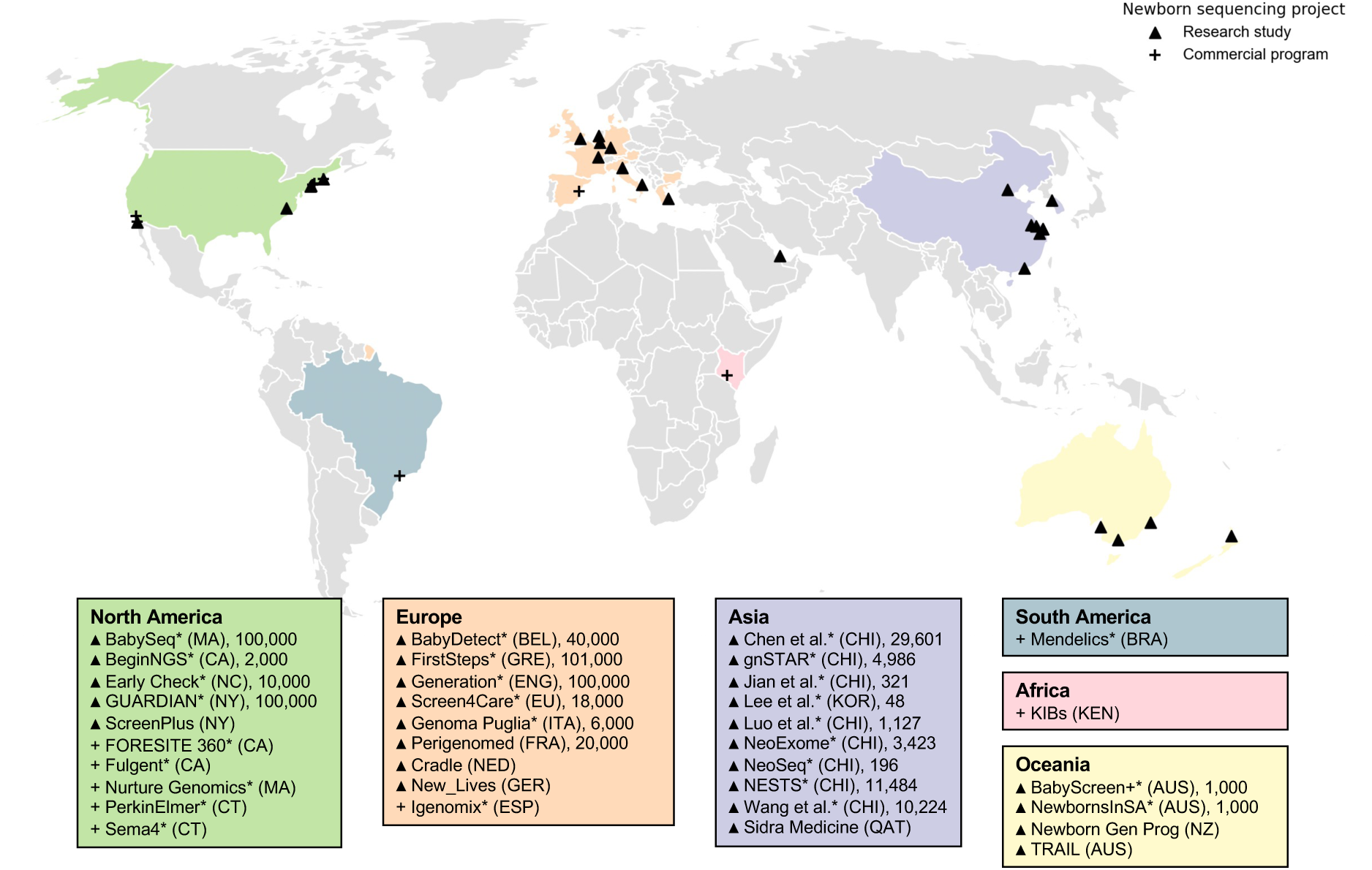
Research and commercial genomic newborn screening (NBSeq) programs. Gene lists from 27 of these programs were included in the analysis (denoted with an asterisk). Intended enrollment sizes are indicated where available.

**Table 1.**
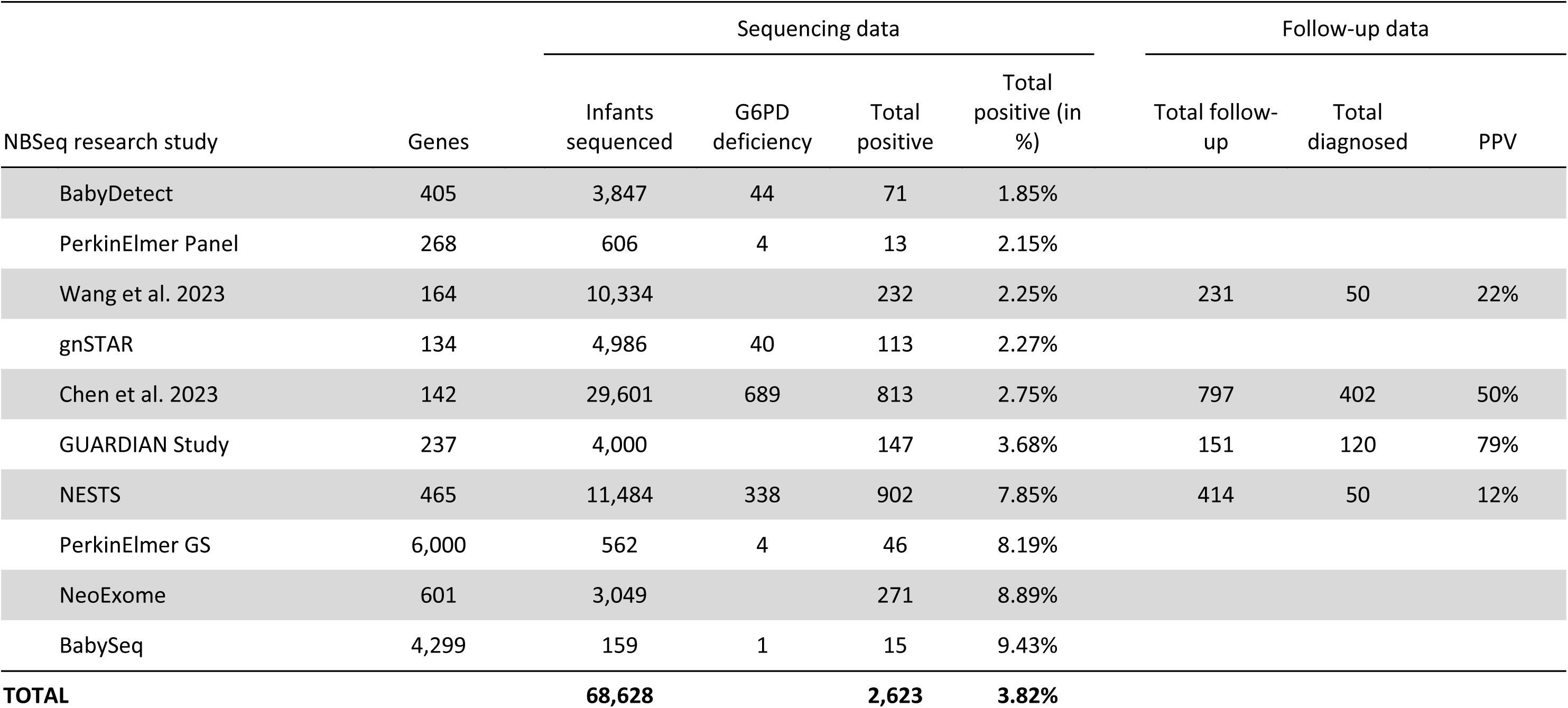
Percentage of positive results from genomic newborn screening research programs. Positive cases of *G6PD,* as well as follow-up data for positively screened infants reported where available. PPV, positive predictive value.

### Compilation of a dataset of gene-disorder characteristics

All obtained gene lists were aggregated, standardizing gene names to HGNC nomenclature and linking each gene to a single disorder using OMIM and ClinGen (Supplementary Methods). A data repository with 25 characteristics for each gene-disorder pair was created, collecting data from five research papers and five existing databases (see Supplementary Methods).^2,10,11,32,51–53^ More information on the source and description of each of the characteristics can be found in the Supplementary Methods and Supplementary Table 3. This data repository as well as the gene list data were made available online in a Github Repository.

### Statistical comparison of gene lists across programs

Descriptive statistics for each gene list, including the length of the list, proportion of genes in each clinical category, and the number of genes associated with RUSP conditions were calculated (Figure 2A, 2B, Supplementary Figure 2, and Supplementary Table 4). To provide information on the concordance across all lists of genes, an UpSet plot, Venn Diagrams, and Jaccard similarity indices were provided (Figure 2C and 2D, Supplementary Figure 3 and 4). Inclusion of high concordance genes, as well as genes associated with core and secondary RUSP conditions, were plotted (Figure 2E, 2F, 2G).

**Figure 2.**
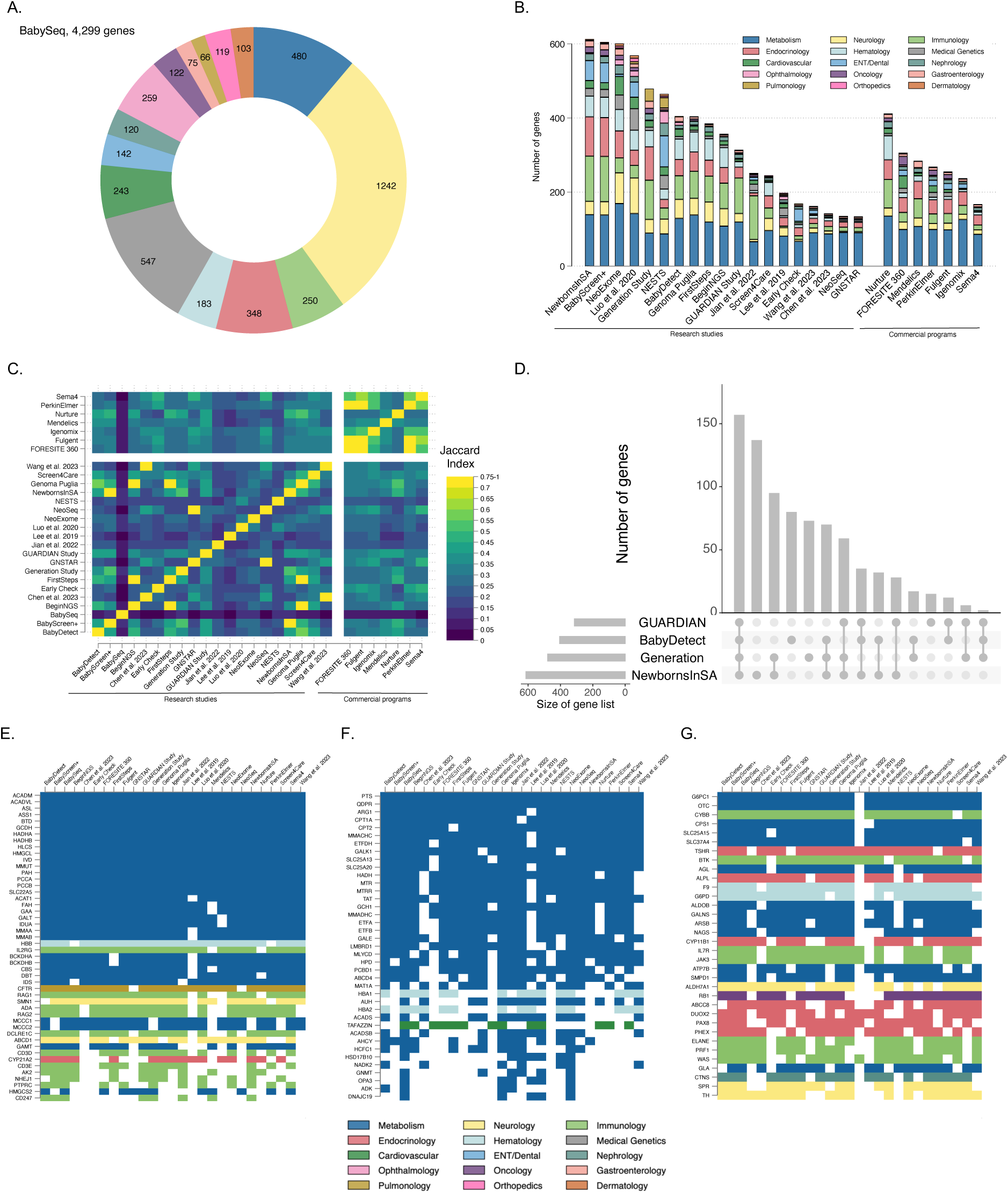
Description, concordance and content of gene lists of genomic newborn screening programs. A. Clinical areas of 4,299 genes included in BabySeq. B. Counts and clinical areas of genes included in 26 research and commercial genomic newborn screening programs (n=4,390). C. Jaccard similarity index, which offers a quantitative comparison of how closely related the gene lists are. D. UpSet plot^65^ of gene lists of 4 large research studies. The matrix below the bar graph represents each individual study and their intersections (n=818). E. Inclusion of genes associated with core Recommended Uniform Screening Panel (RUSP) conditions. The x-axis is each genomic newborn screening program and y-axis are individual genes; the corresponding cell is colored if the gene is included on a given list. F. Inclusion of genes associated with secondary RUSP conditions. G. Inclusion of genes on 20 lists or more that are not associated with RUSP conditions.

A linear regression model was used to identify factors associated with inclusion in multiple gene lists. Two types of regressions were performed: regressions in which the outcome variable is the proportion of gene list inclusion *across all NBSeq programs* (Figure 3A, Supplementary Table 5 and 6) and regressions in which the outcome variable is the inclusion of a gene for each *individual* study (Figure 3B and Supplementary Table 7).

**Figure 3.**
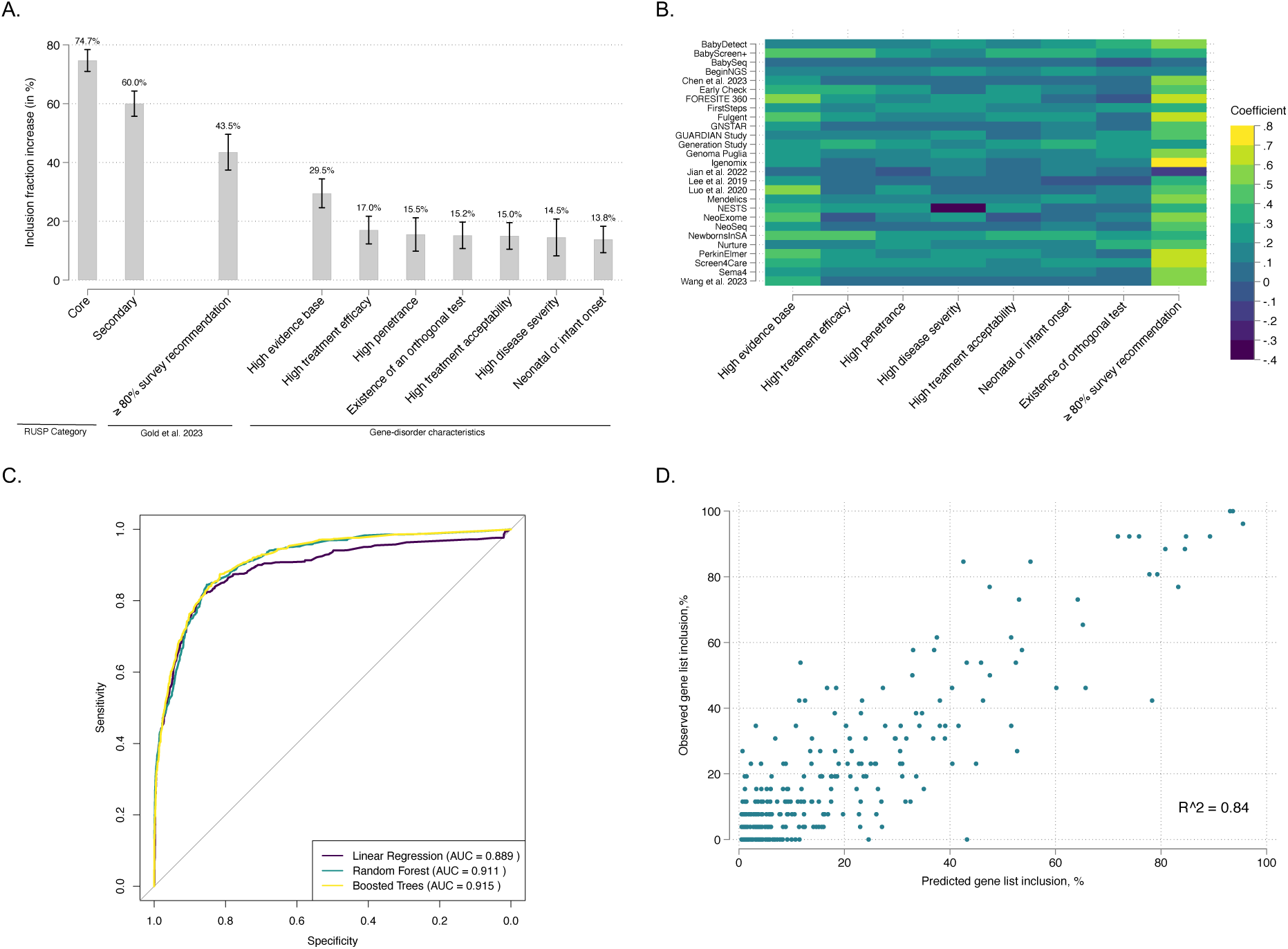
Determinants and prediction model of gene inclusion in genomic newborn screening. In a and b, RUSP category (n=4,474), survey recommendation and orthogonal test (n=649), evidence base, efficacy, penetrance, disease severity, treatment acceptability and neonatal or infant onset (n=749). ROC, Receiver Operating Characteristic; AUC, Area Under the Curve. A. Regression coefficients (and confidence intervals) associated with various gene and disease characteristics predicting inclusion across gene lists. B. Heat map with regression coefficients associated with gene and disease characteristics for each individual genomic newborn screening program. C. ROC curves for three prediction models in the hold-out test set (n=895 genes). D. Scatter plot of predicted versus observed gene list inclusion, showing the fit of the boosted trees model on the 20% hold-out set (n=895 genes).

### Development of a machine learning model

We developed a machine learning prediction model to prioritize genes for population-wide NBSeq programs. Out of 25 potential gene-disorder characteristics, we selected 13 characteristics as predictors in our model: the RUSP category, clinical area, evidence base, severity of the disorder, the treatment efficacy, penetrance, treatment acceptability, age of onset, existence of an orthogonal test, the recommendation score, inheritance, prevalence, and the ClinGen Disease Validity. The remaining 12 gene and disease characteristics were excluded due to a high amount of missing data. For example, the ClinGen actionability scores were not used due to their availability for only 242 genes. Additionally, when characteristics from different sources described similar concepts, we selected the characteristic that includes data on the most genes.

Three machine learning models were compared: linear regression, random forest and boosted trees. We randomly split the gene list data into a 80% training set (n=91,312, 80% of all 114,140 potential instances of 4,390 genes included on a gene list across 26 NBSeq programs) and a 20% test set to estimate prediction power of the model. The boosted trees model, which had the highest area under the curve (AUC), was then selected to generate predictions for all 4,390 genes, providing a ranked list of genes (Supplementary Table 8).

## Results

### Overview of NBSeq programs

Of the 35 research and commercial programs identified, 10 were located in North America, 10 in Asia, nine in Europe, four in Australia and New Zealand, one in South America and one in Africa (Figure 1, Supplementary Table 1). The research programs anticipate a combined total sample size of 560,410 infants, with the intended enrollment in each study varying from 48 to 101,000 infants.^43,54^ Nine NBSeq research programs have published or presented the screening results from a collective total of 68,628 infants by September 2024 (Table 1, Supplementary Methods).

The percentage of positive screening results ranged from 1.85% in BabyDetect (3,847 infants screened for 405 genes) to 9.43% in BabySeq (159 infants screened for 4,299 genes), with an average of 3.82% positive results across 68,628 infants. There was a significant positive correlation between the percentage of positive screening results in a program and the number of genes they screened (pearson correlation coefficient of 0.653, p=0.041). A majority of the collective 1,937 positive screening results across seven studies for which detailed results were available were due to variants in *G6PD* (56.8%).

Four studies reported the clinical outcomes of infants who had undergone NBSeq, allowing for the calculation of these studies’ positive predictive value (PPV), which varied from 12% to 79%^49,50^ with an average across studies of 41% (39% when weighted by sample size).

### Description of gene lists across NBSeq programs

A total of 23 programs have published or made available criteria for selecting genes and disorders for screening (Supplementary Table 2). The number of genes included in 27 programs ranged from 134 to 4,299 (median=306) (Figure 2A, 2B). A total of 4,390 genes (out of a total of 4,966 genes associated with human disease)^55^ were included across at least one of the 27 gene lists (Supplementary Table 4). Of these, 4,033 genes (91.8%) were associated with a phenotype in the OMIM database. Collectively, most genes were linked to inherited metabolic disorders (IMDs) (25.4%), neurologic (15.5%), and immunologic (11.9%) disorders (Figure 2A, 2B).

### Discordance among gene lists used in NBSeq programs

A pairwise Jaccard Index indicated that, aside from those from commercial laboratories, most pairs of gene lists from NBSeq research programs were highly discrepant (Figure 2C and 2D, Supplementary Figure 2, 3). Of the 4,390 genes included in at least one NBSeq program, the vast majority were included by only a small number of NBSeq programs: 4,089 genes (93%) were included by 10 or fewer programs and 3,793 (87%) genes were included by five or fewer programs (Supplementary Figure 1).

### Genes with high concordance across NBSeq programs

Despite this variability across gene lists, we found 74 genes (1.7% of 4,390) that were included by over 80% (22 of 27) of NBSeq programs (Supplementary Figure 1). Of these genes, 58 were associated with diseases on the US Recommended Uniform Screening Panel (RUSP) (Figure 2E, 2F). A total of 34 genes not linked to disorders on the RUSP appeared on 20 or more lists (Figure 2G).

### Predictors of gene inclusion across NBSeq programs

Genes associated with core or secondary disorders on the RUSP were significantly more likely to be included in NBSeq programs (regression coefficient 74.7%, 95% confidence interval (CI): 0.710-0.784, p<0.01; regression coefficient 60.0%, 95% CI: 0.557-0.643, p<0.01, Figure 3A, Supplementary Table 5). Additionally, genes that were recommended for inclusion in newborn screening by 80% or more rare disease experts in a recent survey^11^ were 43.5% (95% CI: 37.4%-49.6%, p<0.01) more likely to be included than genes that were recommended by fewer experts (Supplementary Figure 5).

A strong predictor of inclusion across NBSeq programs was the ASQM evidence base, previously defined as a metric measuring a combination of gene-disease validity, published descriptions of the natural history of disease, and the availability of practice guidelines for disease diagnosis and management.^51^ Genes with the highest evidence base were 29.5% more likely (95% CI: 24.6%-34.4%, p<0.01) to be included in NBSeq programs than those with less available evidence.

Other characteristics associated with inclusion across NBSeq programs were high efficacy of disease treatment (17.0%, 95% CI: 12.3%-21.7%, p<0.01), high penetrance (15.5%, 95% CI: 9.8%-21.2%, p<0.01), neonatal- or infantile-onset (15.2%, 95% CI: 10.7%-19.7%, p<0.01), high disease severity (14.5%, 95% CI: 8.2%-20.8%, p<0.01), high acceptability of treatment (with regard to the burdens and risks placed on the individual) (15.0%, 95% CI: 10.5%-19.5%, p<0.01), and the existence of an non-molecular test that could be used to confirm the diagnosis (13.8%, 95% CI: 9.3%-18.3%, p<0.01). There was variability in the importance of different characteristics across programs, but programs adhered positive value to all these characteristics with few exceptions (Figure 3B and Supplementary Table 7).

### Measuring evolving knowledge about genes and diseases

We conducted a multivariate regression analysis to predict how changes in specific variables, such as treatability and evidence base, would individually influence the overall regression (Supplementary Table 6). Notably, the introduction of a new, highly acceptable treatment for a disorder with no previous treatment is associated with an increase in the likelihood of inclusion in NBSeq programs by 9.7% (95% CI: 0.1%-19.3%, p<0.05). Similarly, improving knowledge related to the natural history of a gene-disorder pair from “none” to “perfect” would increase the likelihood of inclusion in NBSeq programs by 14.8% (95% CI: 4.8%-24.8%, p<0.01).

### Machine learning prediction model

Of three machine learning methods, the boosted trees model demonstrated the highest accuracy in the hold-out test set, with an area under the curve (AUC) of 0.915 and R-squared of 80% (n=22,828, 20%) (Figure 3C, 3D). The relative importance of all variables in the boosted trees model was highest for characteristics such as the proportion of experts who recommended inclusion of the gene in NBSeq on a recent survey,^11^ RUSP classification, and disease prevalence, confirming the results from the regression analysis (Supplementary Figure 8).

We used the boosted trees model to predict the observed inclusion of genes across all NBSeq programs, resulting in a list of all genes that had appeared in any NBSeq program, ranked by their predicted inclusion probabilities. Given that most genes associated with conditions on the RUSP were highly ranked, a list with these genes redacted was considered to be more illustrative of the novel capabilities of the model (Table 2). This analysis identified five genes *(PTPRC, ACADSB, NHEJ1, CYBA, and GRHPR)* that, despite being highly ranked by the model, were only included in a low proportion of NBSeq programs. To address missing data for some genes that were included across multiple NBSeq programs, we also created a second ranked list that combines the rankings generated by our machine learning model with the proportion of NBSeq programs in which each gene was observed with equal weights (Supplementary Table 8). By integrating these two sources of information, this hybrid list leverages the most comprehensive evidence available to prioritize genes for potential implementation in public health programs.

**Table 2.**
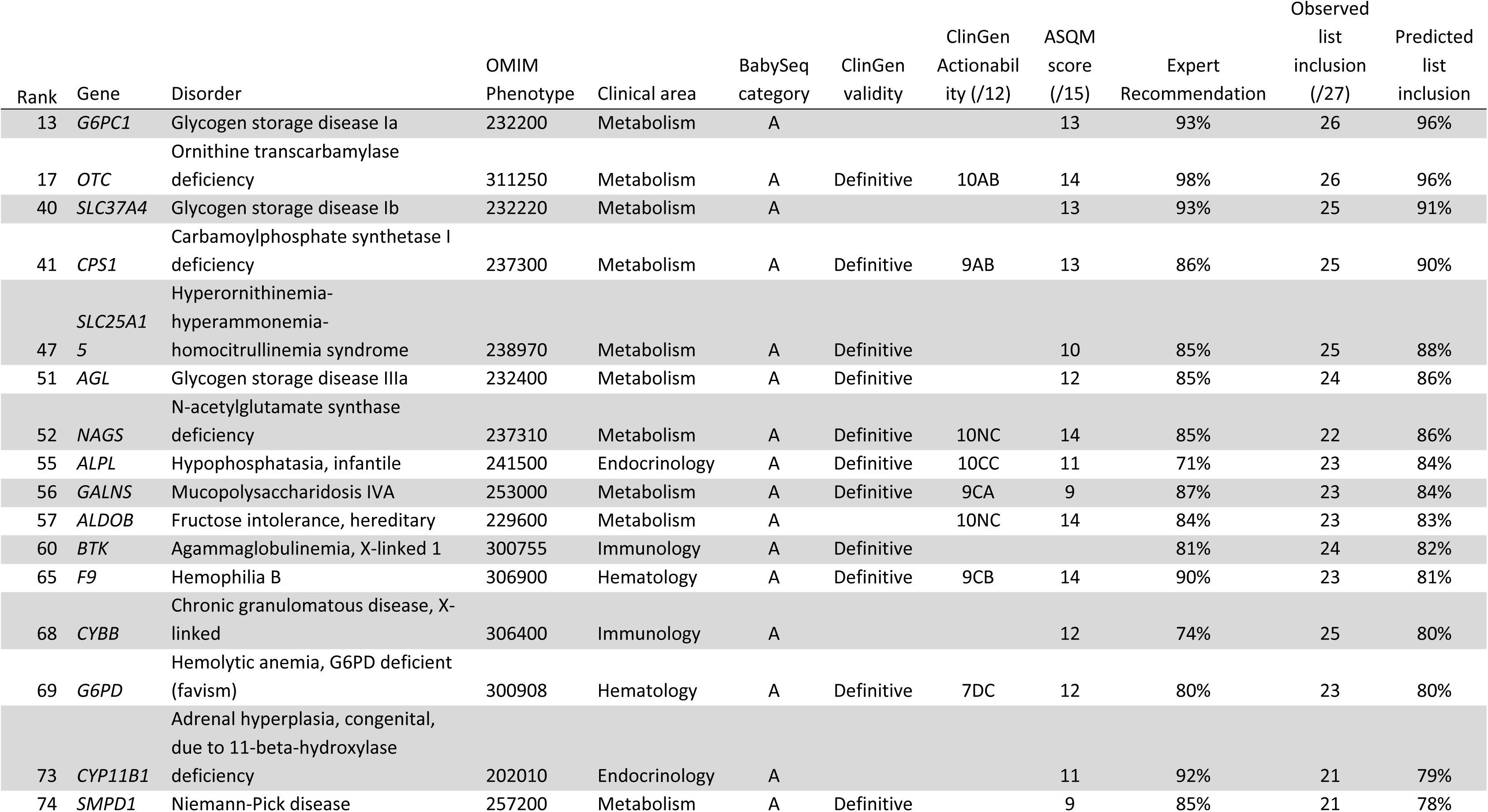
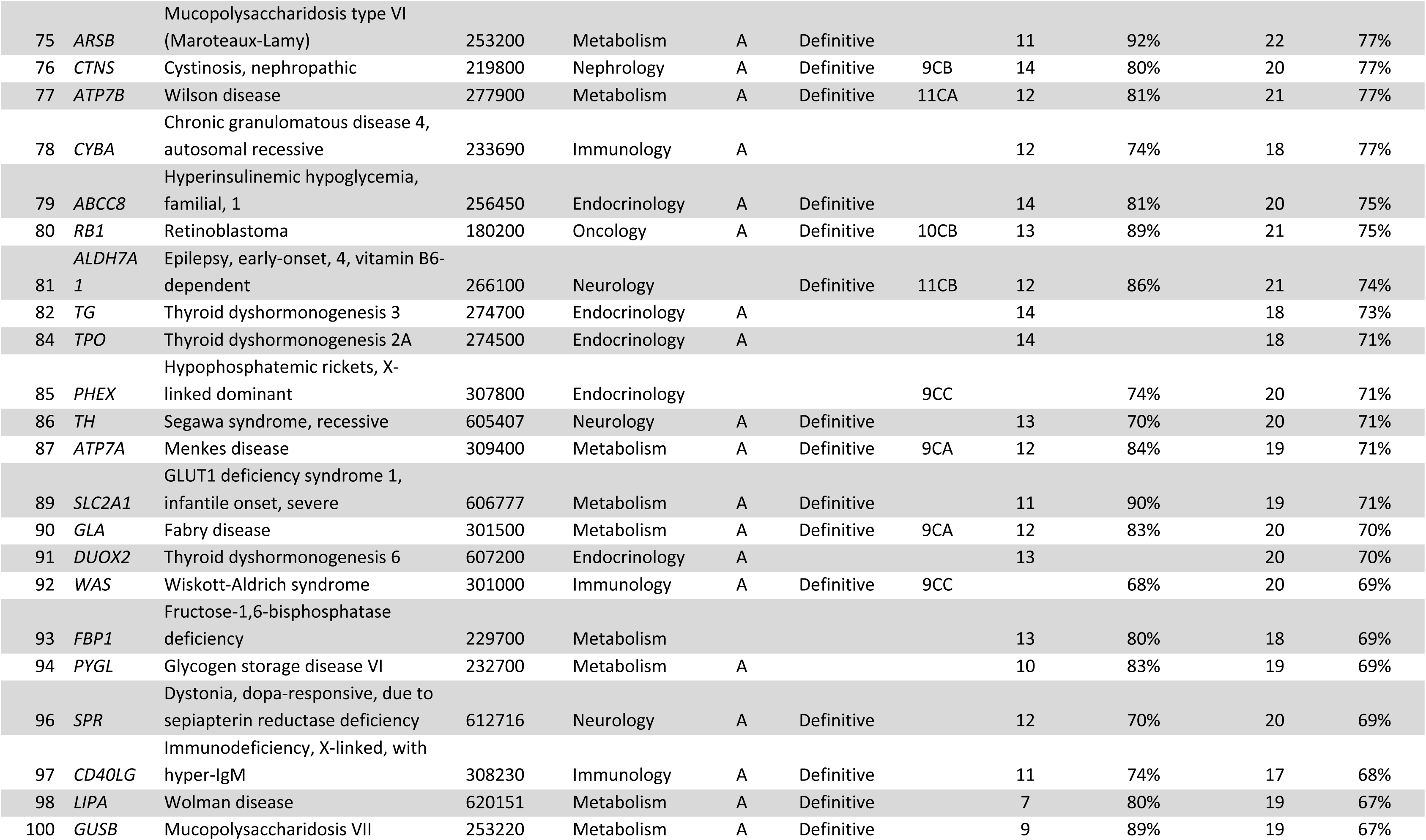
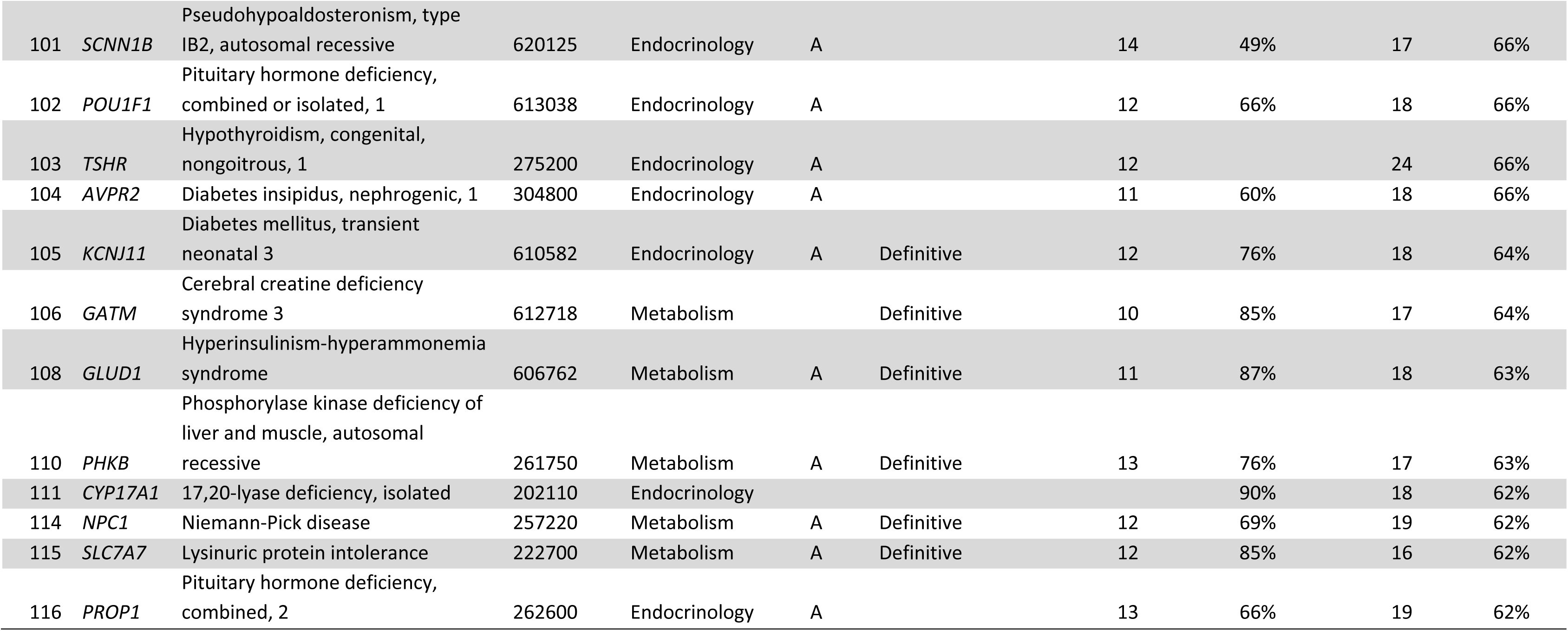
List of 50 genes with highest predicted inclusion across NBSeq programs, excluding genes on the Recommend Uniform Screening Panel (RUSP). ASQM, Age-Based Semi Quantitative Metric.

## Discussion

NBSeq is a rapidly advancing area of global research exploring the impacts of early diagnosis for infants at risk for genetic disorders. With positive screening results in 1.85% to 9.43% of infants and a higher average PPV than some traditional newborn screening techniques,^49,56^ findings from NBSeq research programs support the premise that this approach could improve early detection rates for a wide range of treatable disorders. However, selecting the appropriate genes for screening is a critical step toward implementing population-wide NBSeq. This decision will have significant implications for at-risk infants, their families, and pediatric healthcare systems.^20,21^ In this study, we compared and collected data on the genes being analyzed by 27 NBSeq programs, then developed a machine learning model to predict the inclusion of a gene across NBSeq programs. By combining this model with observed data from NBSeq programs, we generated a ranked list of genes that offers a data-driven approach to prioritizing genetic disorders for public health programs looking to incorporate NBSeq into their screening strategies.

Similar to the findings of smaller studies,^28,29^ our comparison of gene lists from 27 NBSeq programs revealed substantial heterogeneity, which we probed using a series of regression models. We found that the importance of individual gene and disease characteristics varied across studies, potentially due to differences in the international prevalence of disorders, the availability of specialists and treatments in different countries and healthcare systems, or the specific goals of individual NBSeq programs. Unexpectedly, 357 genes with no disease association on OMIM and 52 with limited or refuted gene-disease validity scores from ClinGen were included by some NBSeq programs, demonstrating variation among programs in willingness to include candidate genes or those with new associations to disease.

Despite variations in the gene lists used by NBSeq programs, many share a common focus on certain clinical areas and specific genes. All programs included a substantial proportion of genes associated with disorders that are on the RUSP, reflecting the potential for genomic sequencing to detect cases missed by traditional newborn screening programs.^57–59^ Of note, genes associated with some disorders on the RUSP, such as 3-methyl-crotonyl-CoA carboxylase deficiency (*MCCC1*, *MCCC2*), were widely included across lists despite not conforming to the historic Wilson-Jungner criteria, due to low penetrance and often mild symptoms.^60^ This suggests that some NBSeq programs have anchored their lists around the RUSP even when the disorders are neither severe nor highly treatable.^61^ Therefore, the observed concordance of a gene across NBSeq programs alone may not be sufficient to evaluate the suitability of a gene for population-wide screening.

Many gene and disease characteristics emerged as highly associated with the inclusion of genes across NBSeq programs, including the strength of published data on the natural history of disease, estimated penetrance, and the effectiveness of the associated treatment. Interestingly, despite the inclusion criteria that several NBSeq programs reported, characteristics such as the age of onset and disease severity were weakly associated with inclusion, possibly due to their subjective nature.

The machine learning model developed in this study identifies the disorders that may be most appropriate for genomic newborn screening, based on 13 characteristics and their inclusion across 27 NBSeq programs. This ranked list, along with the preferences of rare disease experts^11^ could be used to prioritize genes for screening, which could then be manually curated by a team of expert reviewers. At this time, the model’s predictions reflect a consensus drawn from NBSeq studies and databases, but in the future, these could be combined with hard-coded gene selection criteria. The model’s flexibility also allows updates based on regional preferences, new data, or emerging therapeutics.

Importantly, this model identified several genes included by only a few NBSeq programs, but which have characteristics that are highly associated with inclusion across programs. For example, although *PTPRC*, a gene associated with severe combined immunodeficiency (SCID), was only included by 12 of 27 NBSeq programs, the model ranked it 113 of 4,390 genes. This is likely because *PTPRC* is associated with a severe immunologic disorder that typically presents in childhood and can be treated with an early hematopoietic stem cell transplant, but is a rare cause of SCID.^62^ This finding highlights the model’s potential to identify genes that may have been overlooked by researchers during the gene selection process.

Our study has several limitations. First, some of the NBSeq programs may have dynamic gene lists that have changed over time. Second, the treatability of various conditions varies based on country. Next, the Jaccard index may exaggerate discrepancies between gene lists of different lengths. For the regression and machine learning models, we consolidated metrics including the ASQM, BabySeq, and ClinGen databases, most of which rely on expert-curated information, such as age of onset, for which definitions vary, and data were missing among the 25 characteristics that we collected. These gaps may automatically reduce the boosted tree model’s predicted inclusion of genes that are not well-characterized, resulting in lower inclusion rates for genes related to disorders that are rare or have limited published evidence. To mitigate the effects of missing data, we designed the model to be easily updated, and provided a ranked gene list that takes into account observed inclusion in addition to our model estimates. The ranked list may suffer from overfitting, given that it includes genes on which the model was trained. Lastly, the model incorporates data from funded research programs and commercial products, which may not align with the goals or constraints of public health newborn screening programs.

In the future, we plan to further develop the dataset that we have created to improve our machine learning model, by incorporating the perspectives of key informants, such as parents^13,14^ and pediatricians. Additional informatics data, such as gene length, gnomAD constraint score,^63^ the ENCODE blacklist,^64^ or the number of associated PubMed publications, will be updated in future versions. We also plan to develop a method to iteratively add genes with newly established gene-disease validity based on recent publications to our dataset. Future research will identify genes that are not currently included in any NBSeq program, but share similar characteristics to those that are highly represented. A knowledge graph, such as PrimeKG,^65^ could be used to find genes that share common molecular pathways, treatments, or symptoms to genes that are currently included in our dataset.

In summary, the growing international interest in genomic newborn screening has prompted important questions about which genes and disorders should be considered for inclusion. Due to the substantial variation in the genes included by 27 NBSeq programs, we developed an evidence-based approach to considering gene selection that draws from a comprehensive data repository encompassing over 4,000 genes. Rather than creating a static list of genes for universal implementation, our dynamic ranking system is adaptable and can be updated as new knowledge about genes, disorders, and therapeutics emerges. This work will support gene selection for both research and public health programs considering the use of population-wide NBSeq.

## Data Availability

All data produced in the present study are available upon reasonable request to the authors.

## Supplementary Methods

### Description of gene lists of genomic newborn screening programs

We obtained gene lists from 27 NBseq programs, including 20 research studies, and seven companies. The 20 research studies are: BabyDetect,^30,66^ BabyScreen+,^28^ BabySeq,^2^ BeginNGS,^32,33^ Chen et al. 2023,^34^ Early Check,^35,48^ FirstSteps, the Generation study, gnSTAR,^36,38^ GUARDIAN study,^42,49,50^ Jian et al. 2022,^37^ Lee et al. 2019,^43^ Luo et al. 2020,^44^ NeoExome,^47^ NeoSeq,^40^ NESTS,^41^ NewbornsInSA, Progetto Genoma Puglia, Screen4Care,^45^ and Wang et al. 2023.^39^ In two studies (GUARDIAN and Early Check), all infants receive testing for a gene list focused on conditions with effective treatments and parents have the option to be tested for an expanded gene list. For both of these studies, we included only the core gene list focused on treatable genetic conditions. Seven lists of genes from commercial firms that offer products related to genomic newborn screening were included: FORESITE 360, Fulgent, Igenomix, Mendelics, Nurture Genomics, PerkinElmer (now Revvity),^46^ and Sema4.^30^ Of note, the Sema4 product is no longer commercially available.

### Rates of positive screening results

A total of nine studies had published or presented results on positive screening results as of September 2024. As studies had different approaches to participant selection, we only reported results of NBSeq programs that screen apparently healthy infants. We excluded results from samples where specifically at-risk infants (such as those in the neonatal intensive care unit) were sequenced. From the Wang *et al* study, only the results on healthy infants were retained.^39^ From the BabySeq study, we included both the healthy and NICU infant sample, as only unanticipated results unconnected with NICU clinical presentations were described for the NICU sample.^5^ We excluded a sample from the NeoEXOME study of neonates that had positive results in conventional NBS.^47^

### Aggregation of gene lists

Gene names were converted to the current nomenclature set forth by the HUGO Gene Nomenclature Committee (HGNC) based on an available online multi-symbol checker.^67^ For purposes of analysis, each gene was linked to one condition. The multistep process for linking genes to a single disorder began by first identifying the phenotypes associated with each gene on Online Inheritance in Man (OMIM).^55^ If only one disease name was associated with the gene on OMIM, a gene-disease pair was formed. If the gene was known to be associated with multiple OMIM disorders, we used the ClinGen gene-disease validity resource to select only the disorder with definitive classification when available.^53^ For genes with more than one disorder with definitive validity or for genes without any disorder with definite validity, one disorder was selected based on the highest number of programs in this study that indicated it as screening target. For example, for *RYR1*, which has a definitive association with both susceptibility to malignant hyperthermia and myopathy,^53^ susceptibility to malignant hyperthermia was selected as the target disorder. Susceptibility to malignant hyperthermia was indicated as a target disease by five of seven programs with disease information available screening for this gene, compared with myopathy which was listed as a target disease by only two of seven programs. A total of 264 genes were not annotated with disorder information from either ClinGen or OMIM databases, possibly because they were candidate genes or had very recently been substantiated as disease genes. These were then manually matched to diseases by search in HGMD (https://www.hgmd.cf.ac.uk/ac/index.php). Two gene names, *GTM* and *CD1*, which could not be linked to HGNC approved gene names, were omitted from the analysis.

### Establishing a data repository of characteristics of genes-disorder pairs

We established a data repository consisting of 25 characteristics for all gene-disorder pairs, sourced from ten different references: five research papers and five existing databases.^2,10,11,32,51–53^ Matching of data from these databases was based on finding an exact match with the gene-disorder pair. Matches were then manually checked for correctness. Three variables (prevalence, clinical area, and RUSP category) were newly constructed for this study and were based on consolidated information from varying sources. Supplementary Table 3 provides an overview of all variables, as well as a description of all metrics and their respective sources.

To determine whether each gene-disease pair was associated with a disorder listed on the United States Recommended Uniform Screening Panel (RUSP), we cross-referenced the genes identified by Owen *et al^33^* with the diseases listed on the RUSP section of the United States Health Resources and Services Administration (HRSA) website (https://www.hrsa.gov/advisory-committees/heritable-disorders/rusp). A gene was considered associated with a RUSP-listed disease if it appeared in the "Cause" section of the corresponding disease-specific HRSA webpage.

Each gene and its associated condition was assigned to one of 12 clinical areas. Although this methodology may not reflect the pleiotropic nature of each disease, the co-authors attempted to identify which pediatric specialist may act as the primary coordinator of an affected child’s care, and which clinical specialty would therefore be most affected by the expansion of NBSeq. The clinical areas included cardiovascular, dermatology, ENT/Dental, endocrinology, gastroenterology, hematology, immunology, medical genetics, metabolism, nephrology, neurology, oncology, ophthalmology, orthopedics, and pulmonology. For 649 disorders, clinical area assignments were based on a previously published paper.^11^ For all other gene-disorder pairs, one medical geneticist or medical student assigned the clinical area (T.M., S.B.), and another medical geneticist or genetic counseling student verified this assignment (S.A., S.B., N.B.G.).

Prevalence estimates for gene-disease pairs were obtained from four sources: Orphanet (https://www.orpha.net/), RX-genes (https://www.rx-genes.com/), and two previously published studies.^11,32^ Gene-disorder links in Orphanet were established through the Orphacode, and only global prevalence estimates were retained. Only prevalence estimates that could be linked to a single gene were withheld. Consolidated prevalence estimates were selected based on the following ordering: RX-genes, Orphanet, Kingsmore *et al* (2022)^32^, and Gold *et al* (2023).^11^ Prevalence data were obtained from these sources for 1,332 gene-disease pairs, covering 30% of the 4,390 genes. Notably, 77.33% of the genes which could be linked to a unique disorder for which a prevalence estimate could be found, had a prevalence of 1 per 1,000,000 or less, highlighting the ultra-rare nature of many disorders included in newborn sequencing programs.

Additional gene and disorder characteristics, including disease penetrance, severity, treatment acceptability and efficacy, age of onset, evidence base (which refers to the level of knowledge about the natural history of the disorder and its treatments), inheritance patterns and the existence of an orthogonal confirmatory non-diagnostic test, were derived from five previously published studies.^2,11,51,52^ When two modes of inheritance were implicated for the same gene and disease in these studies, such as for *MYO6*, a cause of non-syndromic deafness, dominant inheritance was selected as it was expected to lead to the most inclusive reporting criteria.

Gene-disorder pairs were also matched with ClinGen (https://clinicalgenome.org/) gene-disease curations, which were evaluated using a standardized approach to assess the strength of evidence linking a gene to a monogenic disease. Additionally, ClinGen clinical pediatric and adult actionability curations were obtained, with the highest actionability score retained when multiple curations were available.^8,52^ For gene-disorder pairs with both pediatric and adult curations, only the pediatric score was retained.

Three existing metrics that address the suitability of genes for newborn sequencing were also included. The age-based semi-quantitative metric (ASQM) score^51,52^ is a metric which assigns a number between 0 and 15 to a gene-disease pair to denote overall appropriateness for newborn screening based on several sub-scores. The BabySeq Category is another metric scored by the BabySeq Project,^2^ where BabySeq Category A is designated as the category of genes most amenable to newborn screening, while Category C is considered less amenable to newborn screening. Finally, we incorporated the proportion of 238 rare disease experts who recommended screening newborns for 649 genes, as determined by an online survey.^11^

### Additional information on the statistical analyses

All statistical analyses were carried out in Stata 18 (College Station, TX), R version 4.3.1 (Vienna, Austria) and Python version 3.11.2 (Python Software Foundation, Beaverton, OR). Jaccard similarity indices were calculated to provide information on the concordance across all lists of genes (Figure 2C and Supplementary Figure 4). The Jaccard index measures the number of genes in the intersection set divided by the number of genes in the union set of two gene lists. The UpSet plot was plotted using the UpSet^68^ library in R (Figure 2D).

Regression coefficients on each characteristic were measured in a separate regression, where the only other control was the RUSP category (Figure 3A, Supplementary Table 5). Regression coefficients were also obtained for each program separately running several regressions for each program, one for each program-characteristic combination, while also controlling for RUSP category (Figure 3B, Supplementary Table 7). Several multivariate regression models were evaluated (Supplementary Table 6). Among them, specification (5), which included the most comprehensive set of independent variables, was used to estimate the effect of the change in a gene characteristic on overall gene list inclusion. However, it is important to note that this analysis identifies associations rather than causal relationships, even with the inclusion of additional controls.

The distribution of genes across several quantitative scores previously designed to assess the overall usefulness of disorders for NBSeq, including the BabySeq Category^2^ and ASQM Score^51,52^ were calculated within each program (Supplementary Table 4), and correlations with overall gene inclusion across NBSeq programs was plotted (Supplementary Figure 6).

### Additional information on the machine learning model

The prediction analysis was implemented in R using linear regression, random forest and boosted trees (glm, randomforest and gbm packages). Our machine learning algorithm was developed to predict a binary variable: whether a gene was included in an NBSeq program’s gene list. We also experimented using the proportion of 27 NBSeq programs that included the gene as an outcome measure, but results were similar. Since the BabySeq project uses an “elective exome approach” that includes nearly all genes associated with human disease, we excluded this list when training the model. The overall dataset used for prediction consisted of 114,140 program-gene observations. We randomly assigned 80% of genes (91,312 observations) to the training set, and 20% to the hold-out test set.

For the 13 gene and disease characteristics included in the model, missing values were handled by adding dummy variables in the regression model and setting the missing predictor to zero. In the random forest and boosted trees models, missing values were set to -1 or labeled as ‘missing’ in the case of categorical variables.

We conducted a grid search using cross-validation to optimize the hyperparameters in the boosted trees model using the caret package. We optimized interaction depth (interaction.depth: 1, 5), number of trees (n.trees: 100, 500), learning rate (shrinkage: 0.01, 0.1), and minimum observations per node (n.minobsinnode: 10, 20). The final model configuration was chosen based on its performance during three-fold cross-validation, aiming to maximize predictive accuracy.

The performance of each model was evaluated using Receiver Operating Characteristic (ROC) curves and the Area Under the Curve (AUC) metric (Figure 3C). We plotted distribution plots, showing the distribution of predictions for all models (Figure 3C, Supplementary Figure 7). Additionally, we plotted a calibration plot, showing the observed versus predicted inclusion probabilities for each gene-disorder pair in the test set (Supplementary Figure 7). We assessed the importance of gene and disease characteristics in the boosted trees model using the gbm package, where importance was measured by the mean decrease in accuracy for random forest and reduction in deviance for boosted trees (Supplementary Figure 8).

## Acknowledgements

This work was supported by the following grants: T32GM007748 (S.B.), R01HG011773 (N.G.), K08HG012811-01 (N.B.G.), TR003201 (N.B.G.), HD077671 (R.C.G.) TR003201 (R.C.G.), and EU-IMI H2020 GRANT 101034427 (A.F., J.K.).

## Ethics declaration

This study did not involve experiments on human participants or animals. All outcome data utilized in this research were aggregated and obtained from previously published sources, which are cited within the manuscript. As no individualized or patient-specific data were used, Institutional Review Board (IRB) or Research Ethics Committee (REC) approval were not required.

## Competing interests

L.M.A., A.J.C. and R.J.T. are employees and shareholders at Illumina Inc. N.G. is co-founder and equity owner of Datavisyn. N.B.G. provides occasional consulting services to RCG Consulting and receives honoraria from Ambry Genetics. R.C.G. has received compensation for advising the following companies: Allelica, Atria, Fabric, Genomic Life and Juniper Genomics; and is co-founder of Genome Medical and Nurture Genomics. B.E.R. and K.L.S. are consultants at Nurture Genomics. L.S. received personal compensation from Zentech and Illumina Inc. P.T. is a co-founder of PlumCare RWE, LLC.

## Author contributions

Conceptualization: L.M.A., D.B., F.B., A.J.C., N.E., A.F., N.B.G., R.C.G., J.K., T.M., B.E.R., K.L.S., L.S., R.J.T. Data curation: S.B., A.J.C., T.M., K.L.S, H.Z. Formal analysis: N.B.G., T.M. Funding acquisition: R.C.G, L.S., P.T. Investigation: S.A., S.B., N.B.G., T.M. Methodology: N.G., N.B.G., R.C.G., T.M. Resources: H.Z. Project administration: T.M. Software: T.M. Supervision: N.B.G., R.C.G., L.S. Visualization: N.G., N.B.G., T.M. Writing-original draft: S.A., N.B.G., T.M. Writing-review & editing: all authors.

## Data and materials availability

All datasets generated and/or analyzed in this study, along with the Stata, R, and Python code necessary to replicate the results, are available at https://github.com/tjminten/gim_genelist.

## Web resources

BabyDetect, https://babydetect.com/en/

BabyScreen+, https://babyscreen.mcri.edu.au/

BabySeq, https://www.genomes2people.org/research/babyseq/

BeginNGS, https://radygenomics.org/begin-ngs-newborn-sequencing/

Early Check, https://earlycheck.org/

FirstSteps, https://www.firststeps-ngs.gr/

FORESITE 360, https://foresite360.com/

Fulgent Genetics, https://www.fulgentgenetics.com/

Genomics England Newborn Genomes Programme, https://www.genomicsengland.co.uk/initiatives/newborns

GUARDIAN Study, https://guardian-study.org/

International Consortium on Newborn Sequencing (ICoNS), https://www.iconseq.org/

Igenomix, https://www.igenomix.eu/

Kenya Bioinformatics Institute, https://www.kibs.co.ke/

Mendelics, https://mendelics.com.br/

NEW_LIVES, https://www.klinikum.uni-heidelberg.de/en/new-lives-genomic-newborn-screening-programs

NewbornsInSA, https://www.wch.sa.gov.au/research/other-research-projects/newbornsinsa-research-study

Nurture Genomics, https://nurturegenomics.com/

Screen4Care, https://screen4care.eu/

ScreenPlus, https://www.einsteinmed.edu/research/screenplus/

PerkinElmer (now Revvity), https://www.revvity.com/be-en/category/newborn-screening

## Supplement A: ICoNS Gene List Contributors

Programs:

BabyDetect, BabyScreen+, BabySeq, BeginNGS, Chen et al. 2023, Early Check, FirstSteps, the Generation study, gnSTAR, GUARDIAN study, Jian et al. 2022, Lee et al. 2019, Luo et al. 2020, NeoExome, NeoSeq, NESTS, NewbornsInSA, Screen4Care, Progetto Genoma Puglia, Wang et al. 2023, FORESITE 360, Fulgent, Igenomix, Nurture Genomics, PerkinElmer (now Revvity), Sema4.

Individuals:

| First name and initials | Last name | Degrees | Affiliation | Program |
| --- | --- | --- | --- | --- |
| Mattia | Gentile |  |  | Progetto Genoma Puglia |
| Paola | Orsini |  |  | Progetto Genoma Puglia |
| Romina | Ficarella |  |  | Progetto Genoma Puglia |
| Maria Luisa | Valente |  |  | Progetto Genoma Puglia |
| Emanuela | Ponzi |  |  | Progetto Genoma Puglia |
| Athina | Ververi | MD, PhD, MSc. PgCert | Papageorgiou General Hospital, Thessaloniki, Greece | FirstSteps |
| Maria | Koutsogianni | MD, MSc. | Institute of Child Health Great Ormond Street University College London. | FirstSteps |
| Huang | Xinwen |  |  | gnSTAR |
| Xiao | Rui |  |  | gnSTAR |
| Zhao | Zhengyan |  |  | gnSTAR |
| Matthew J. | Pelo |  |  | FORESITE 360 by Fore Genomics |
| Jovanka | King | PhD<br>FRACP<br>FRCPA<br>BMBS<br>(Hons)<br>BPod | Genetics and Molecular Pathology, SA Pathology, Adelaide SA 5000, Australia<br>Adelaide Medical School, The University of Adelaide, Adelaide SA 5000, Australia<br>The Women's and Children's Health Network, Adelaide SA 5000, Australia | NewbornsInSA |
| Carol | Siu | MBBS,<br>PhD,<br>FRCPA,<br>FRCPath | Genetics and Molecular Pathology, SA Pathology, Adelaide SA 5000, Australia<br>Adelaide Medical School, The University of Adelaide, Adelaide SA 5000, Australia | NewbornsInSA |
| Karin | Kassahn | PhD,<br>FFSc(RC<br>PA) | Genetics and Molecular Pathology, SA Pathology, Adelaide SA 5000, Australia<br>Adelaide Medical School, The University of Adelaide, Adelaide SA 5000, Australia | NewbornsInSA |
| Sansen | Stefaan |  | Sanofi | Screen4Care |
| Enrico | Bertini |  | Paediatric Hospital Bambino Gesù Rome | Screen4Care |
| Aldona | Zygmunt |  | Pfizer | Screen4Care |

## Supplement B: International Consortium on Newborn Sequencing (ICoNS) authors

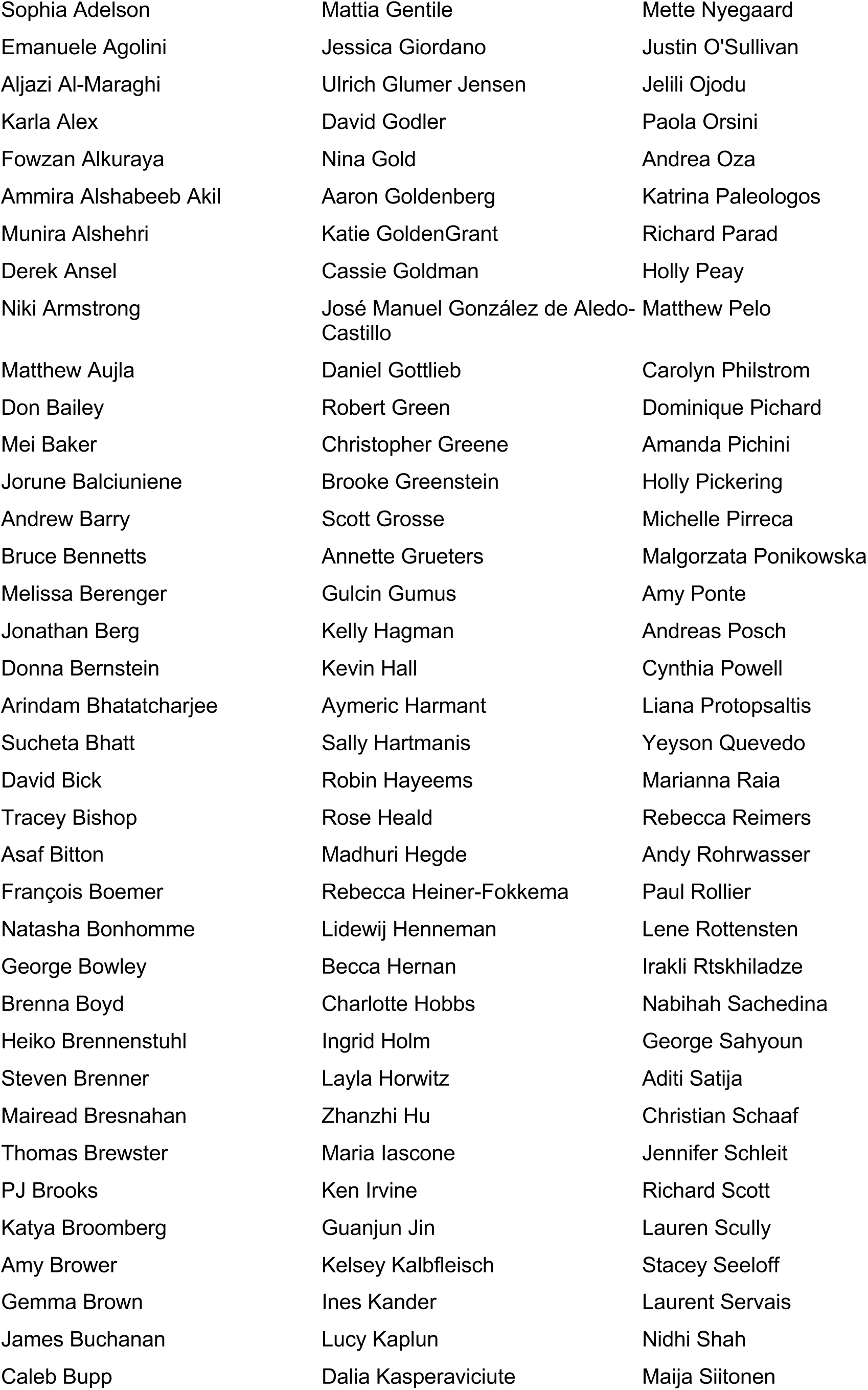
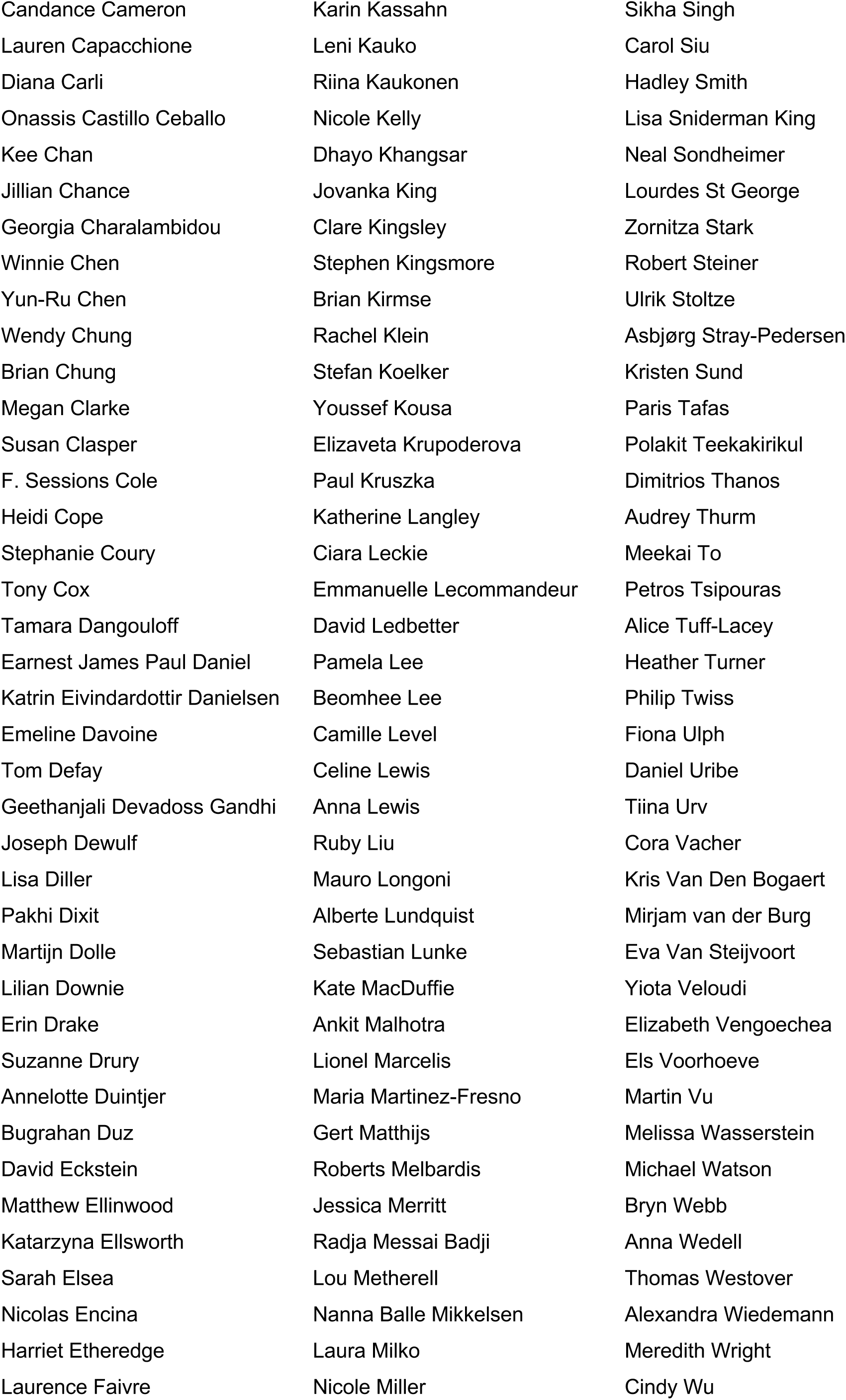
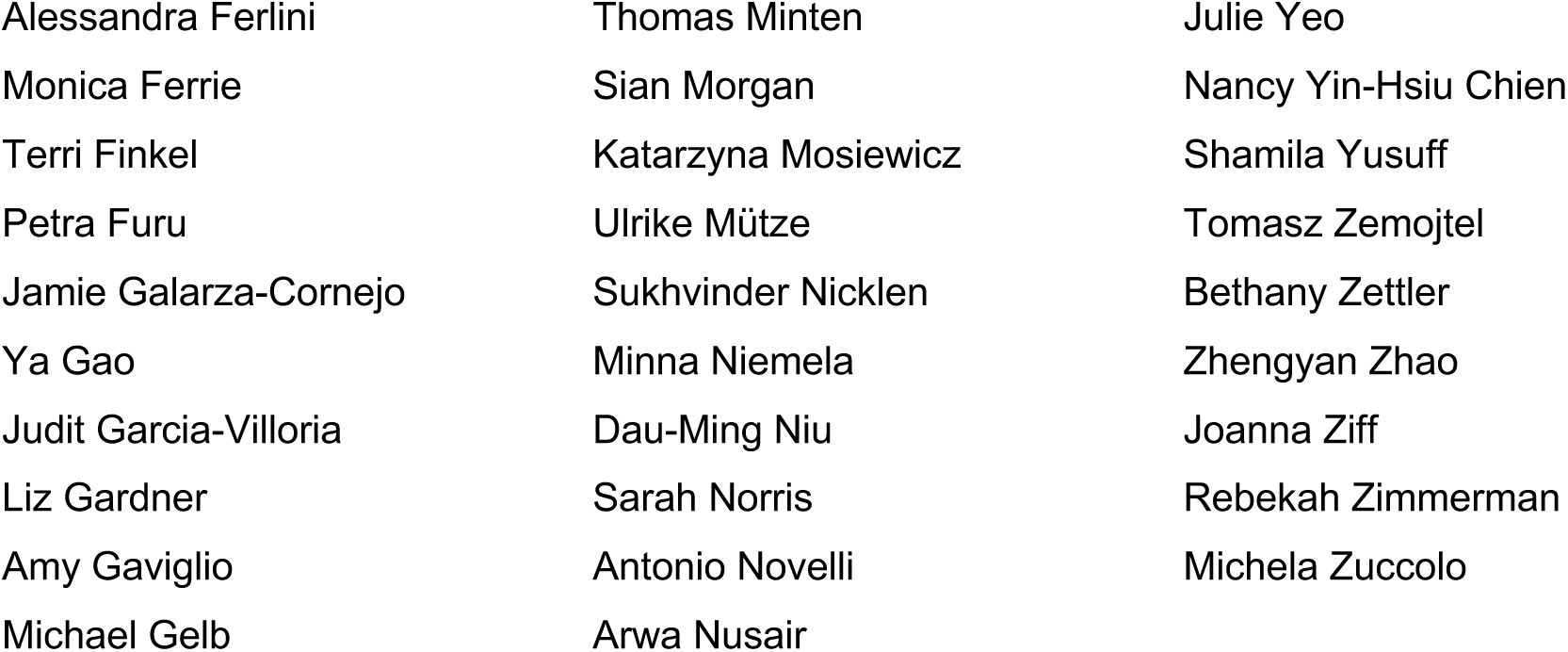

## Supplementary Material

**Supplementary Figure 1.**
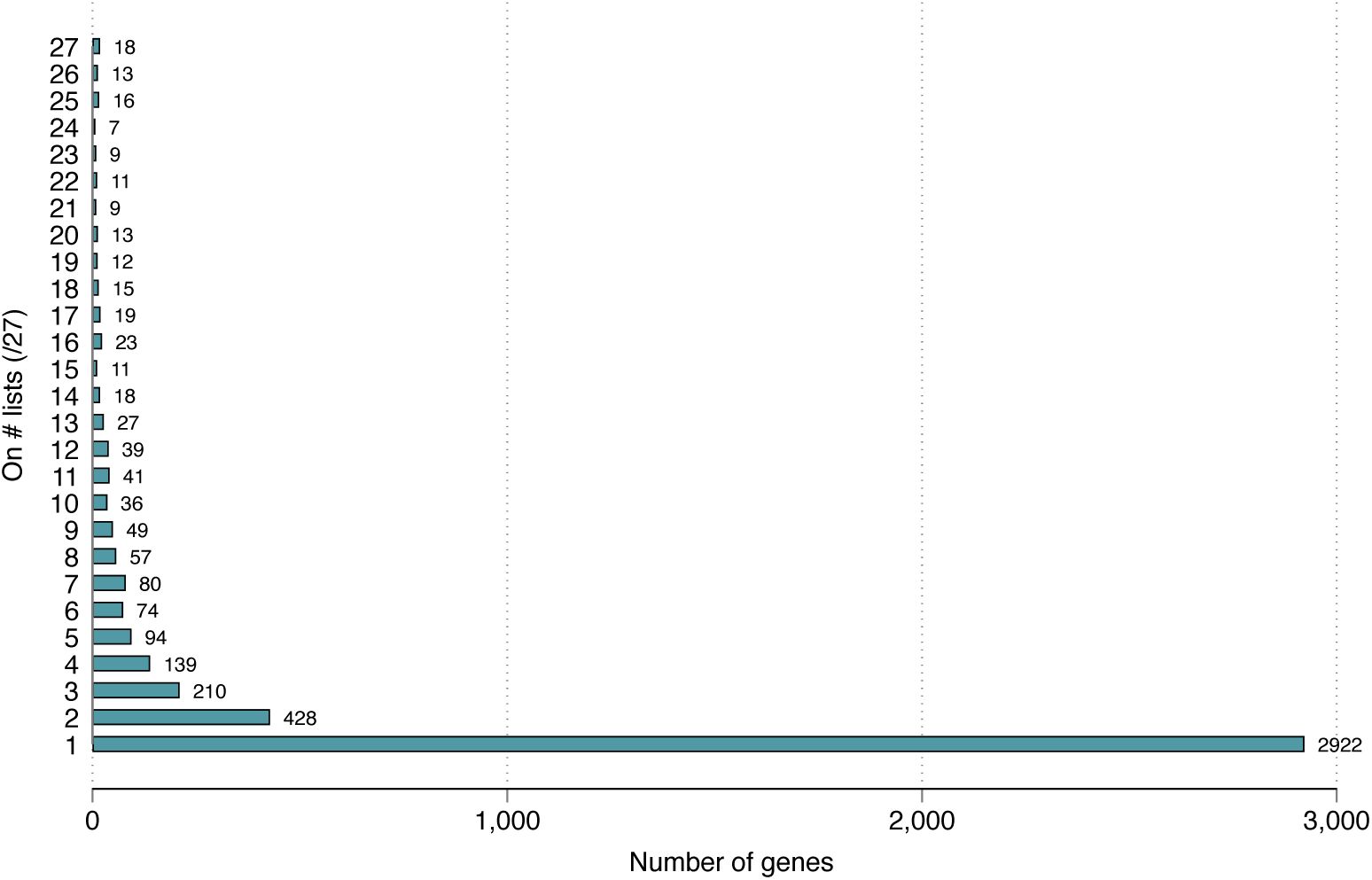
Gene inclusion distribution. **Notes:** The distribution of gene list inclusion for all gene-disorder pairs included in any of the NBSeq programs (n=4,390).

**Supplementary Figure 2.**
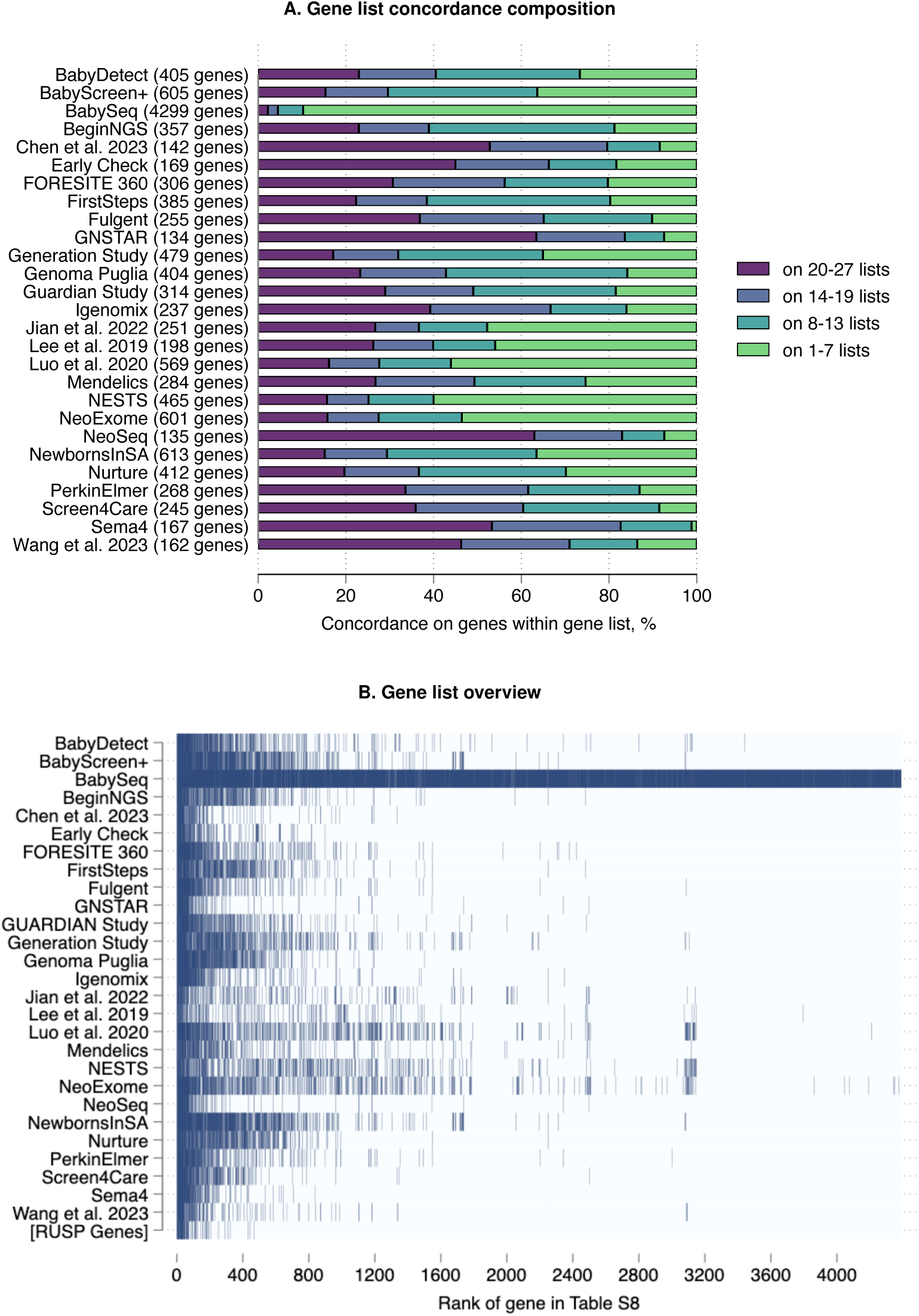
Gene list concordance. **Notes:** (A) A bar chart displaying the distribution of the number of programs that include genes from each program’s gene list, highlighting the overall concordance within each list. (B) A gene map visualizing the presence of 4,390 genes across 27 NBSeq studies. On the x-axis, all 4,390 genes are listed, while the y-axis represents the 27 studies, and the RUSP. Each colored cell indicates that a gene is included in a given study’s list. RUSP, Recommended Uniform Screening Panel.

**Supplementary Figure 3.**
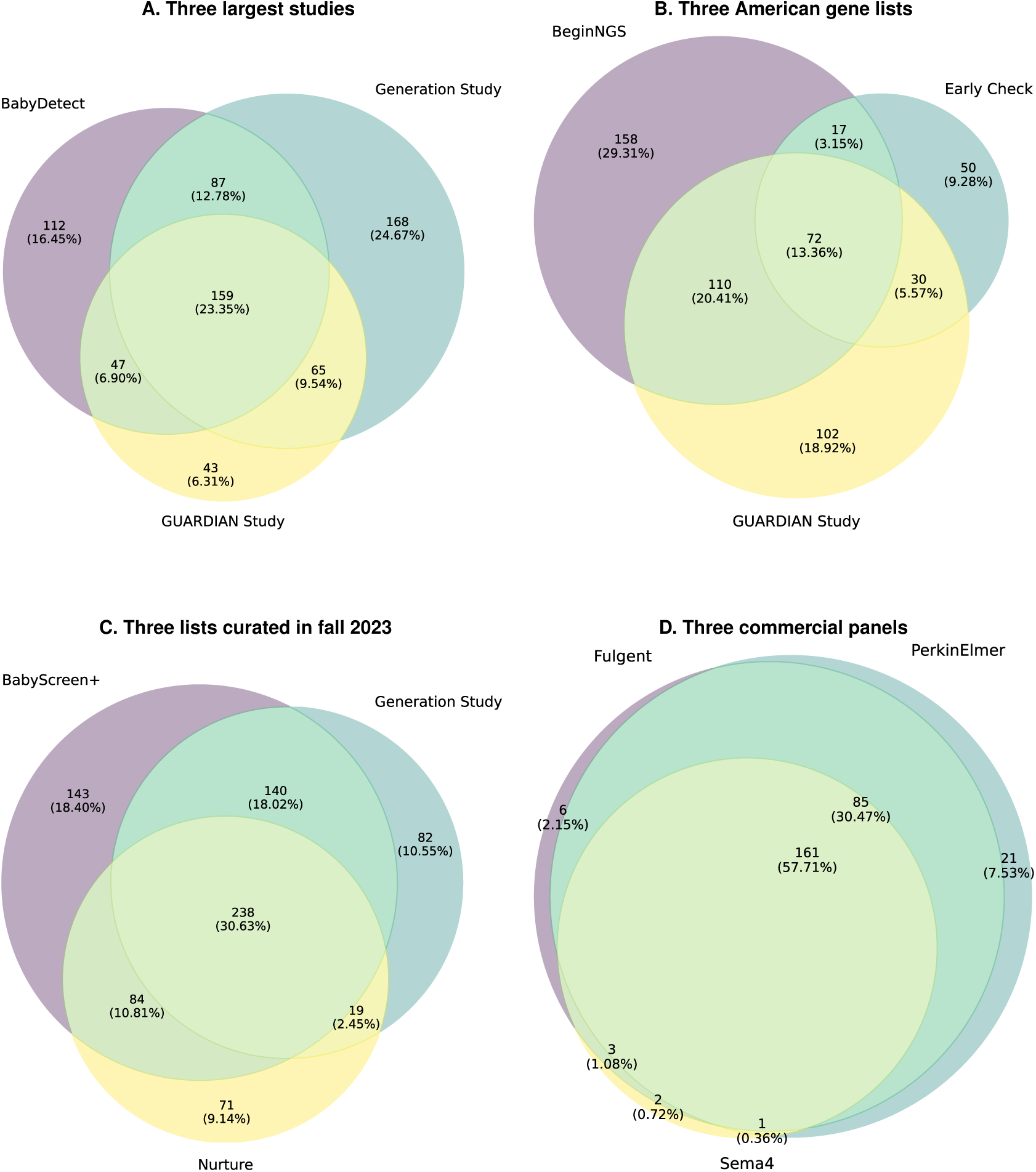
Gene list venn diagrams. **Notes:** Venn diagrams illustrating the overlap between gene lists from different studies. The size of each circle scales proportionally to reflect the number of genes in each study. Within each overlapping region, the numbers indicate the exact count of genes, with the corresponding percentages representing their proportion relative to the total combined gene count across all three studies.

**Supplementary Figure 4.**
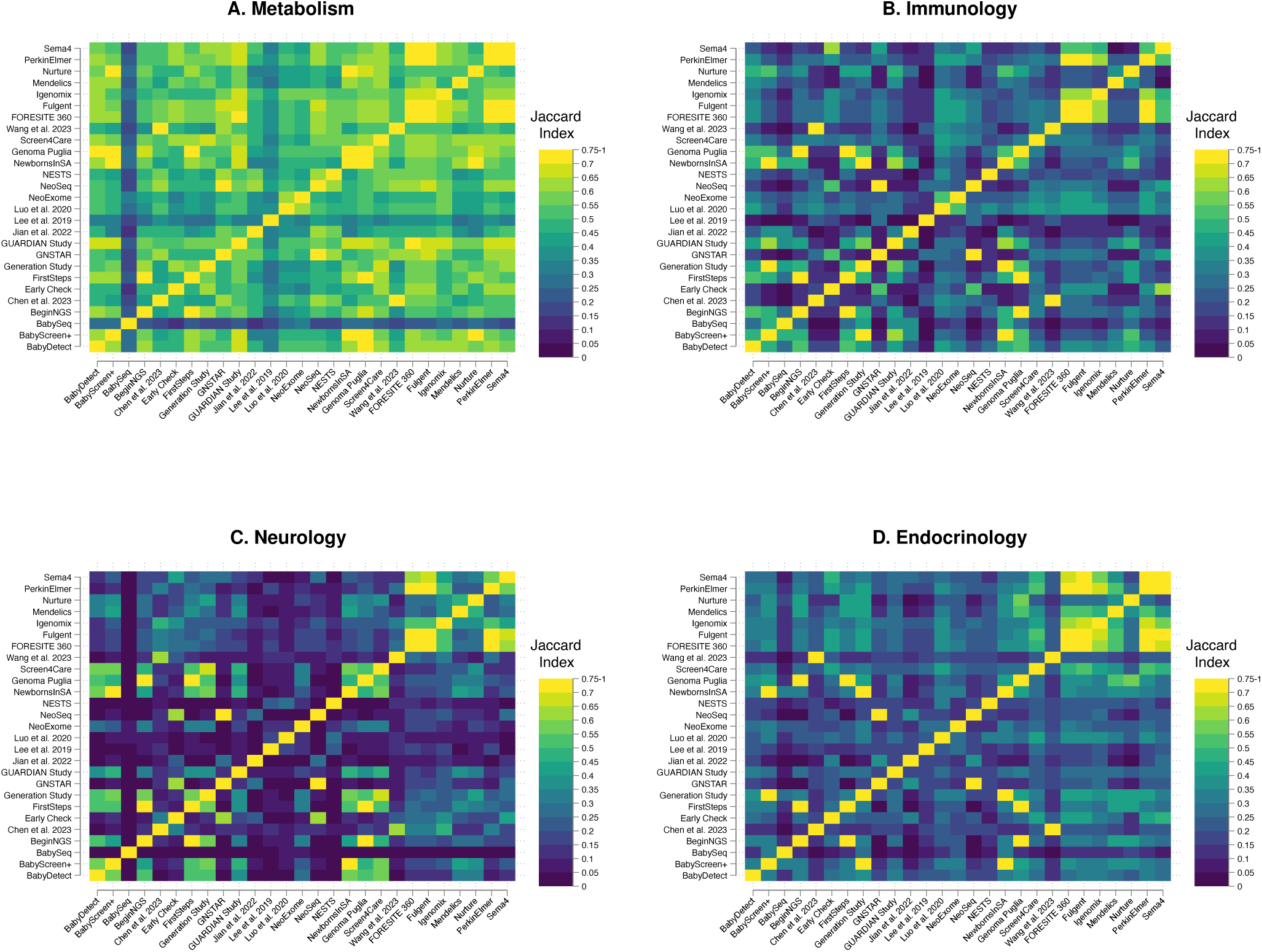
Jaccard heat plots by clinical area. **Notes:** Jaccard similarity indices for all pairs NBSeq programs, looking only at genes associated with (A) metabolic (n=492), (B) immunologic (n=274), (C) neurologic (n=1,251), and (D) endocrinologic disorders (n=352). This index measures the degree of overlap between gene lists by comparing the intersection and union of the sets (see Methods). The resulting indices offer a quantitative comparison of how closely related the gene lists are for these specific types of disorders.

**Supplementary Figure 5.**
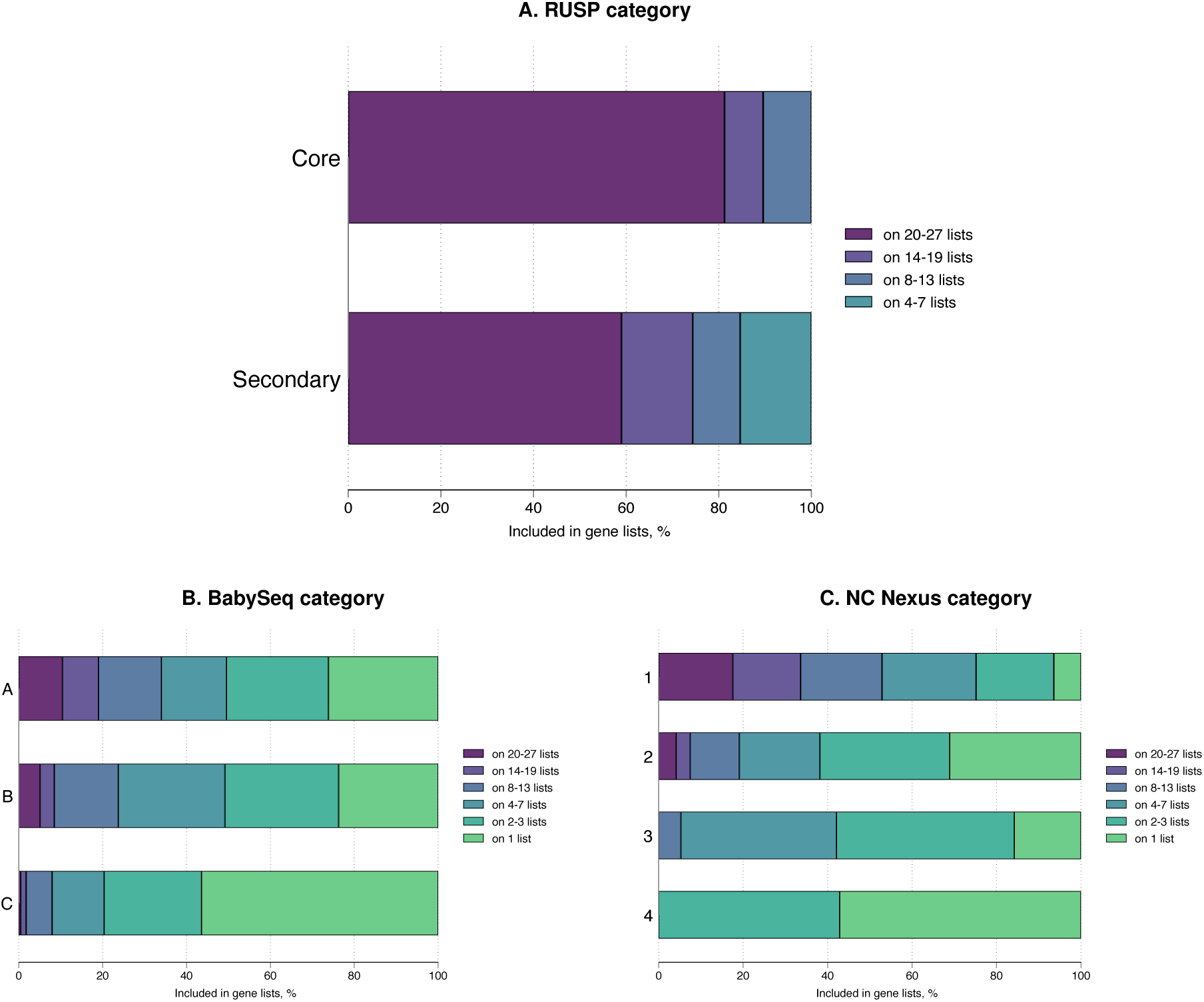
Correlation of gene list inclusion and different categories. **Notes:** Bar charts showing the distribution of gene list inclusion across different categories. The color-coded bars indicate the number of lists each gene is included in. (A) RUSP Core (n=48) and Secondary (n=39). BabySeq Category A (n=826), B (n=59), and C (n=452). NC Nexus Category 1 (n=467), 2 (n=241), 3 (n=19), 4 (n=14). RUSP, Recommended Uniform Screening Panel.

**Supplementary Figure 6.**
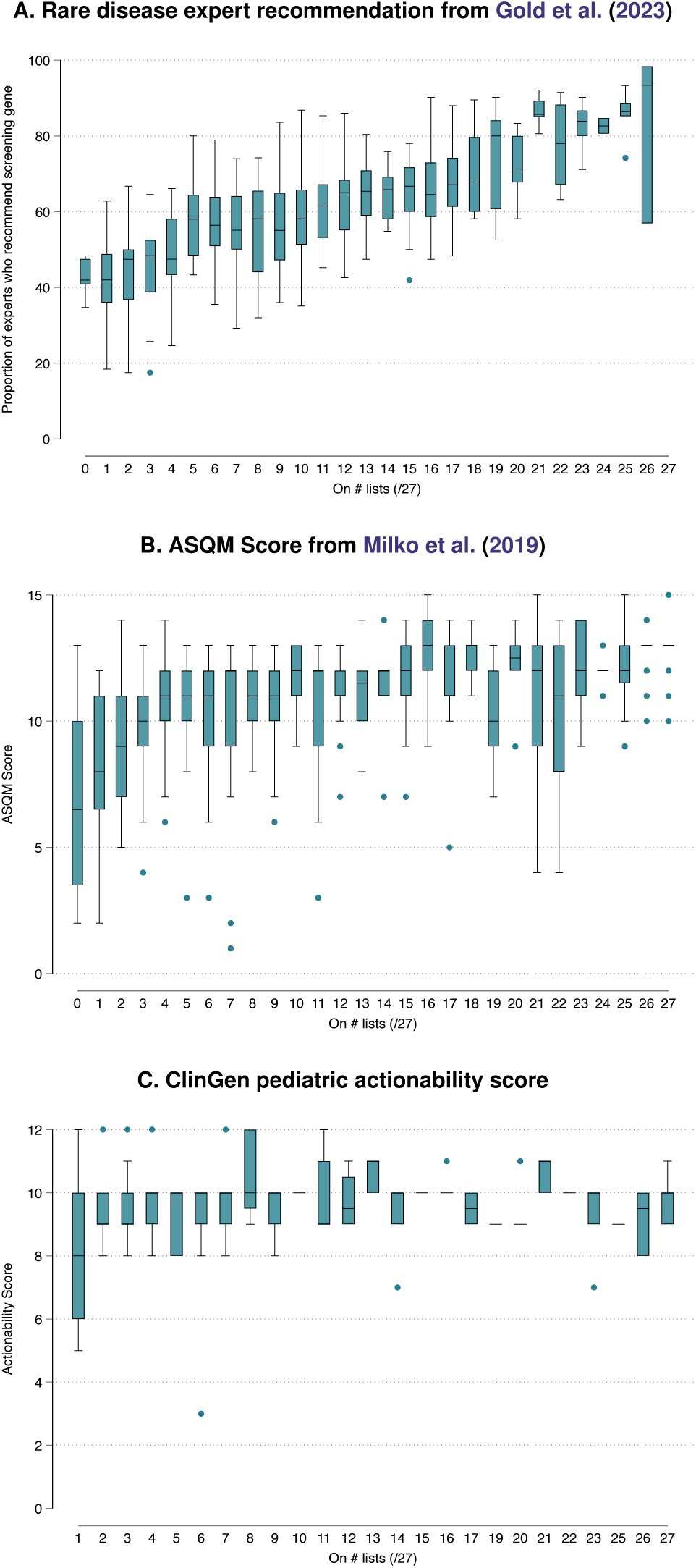
Correlation of gene list inclusion and previously published metrics. **Notes:** (A) Box plot showing the distribution of expert recommendations for gene screening across concordance levels, based on Gold et al. (2023) (n=649). (B) Box plot of ASQM scores for genes, from Milko et al. (2019), across concordance levels (n=747). (C) Box plot of ClinGen pediatric actionability scores across concordance levels (n=135). Each plot displays the median (center line), interquartile range (box), and full range (whiskers) within each group. ASQM, Age-Based Semi Quantitative Metric.

**Supplementary Figure 7.**
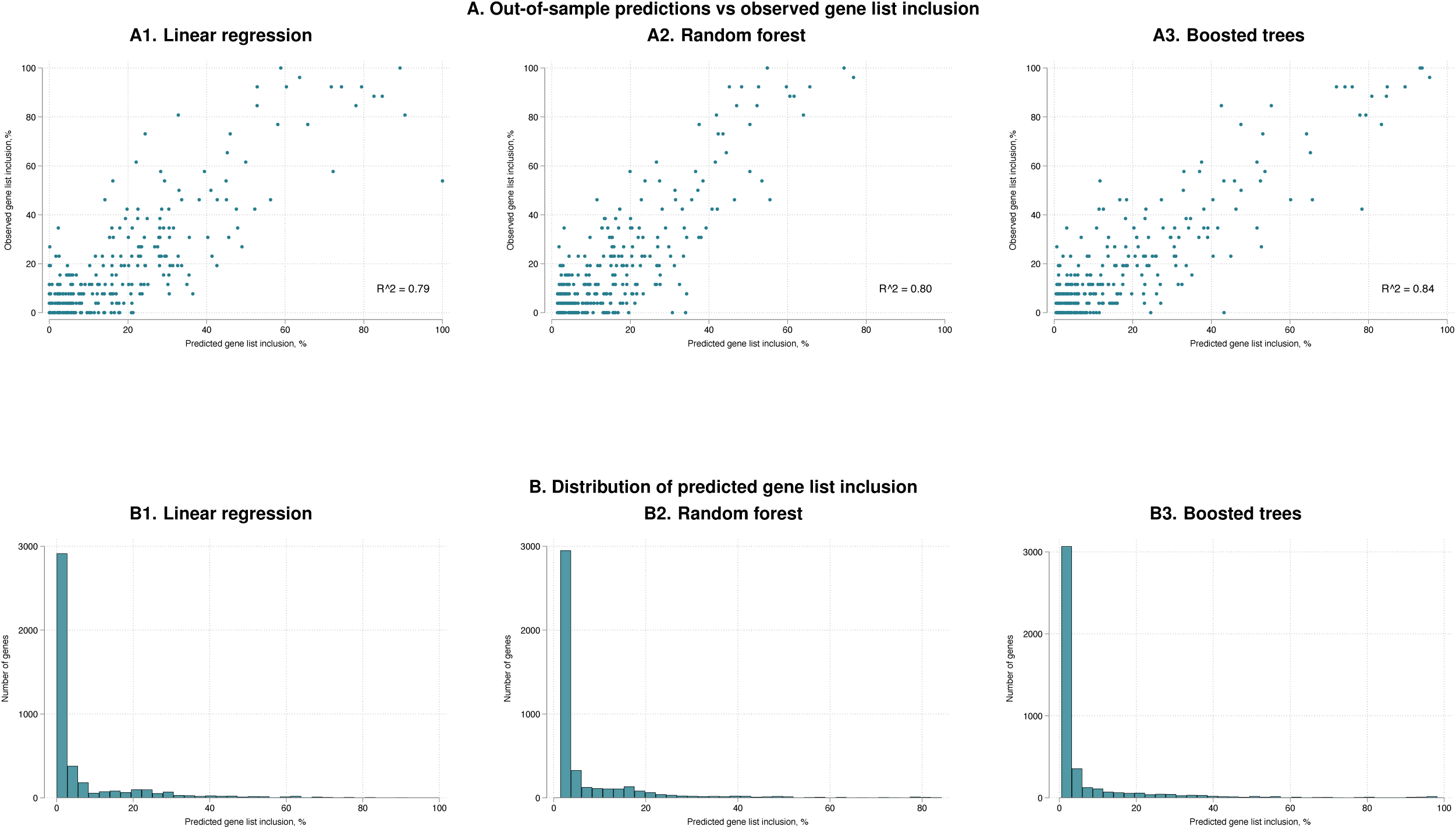
Prediction model graphs. **Notes:** (A) Scatter plots of the 20% hold-out test set, plotting predicted versus observed gene list inclusion. Perfect predictions would align along a 45-degree line. (B) Histogram displaying the distribution of predicted gene list inclusion for 4,390 genes.

**Supplementary Figure 8.**
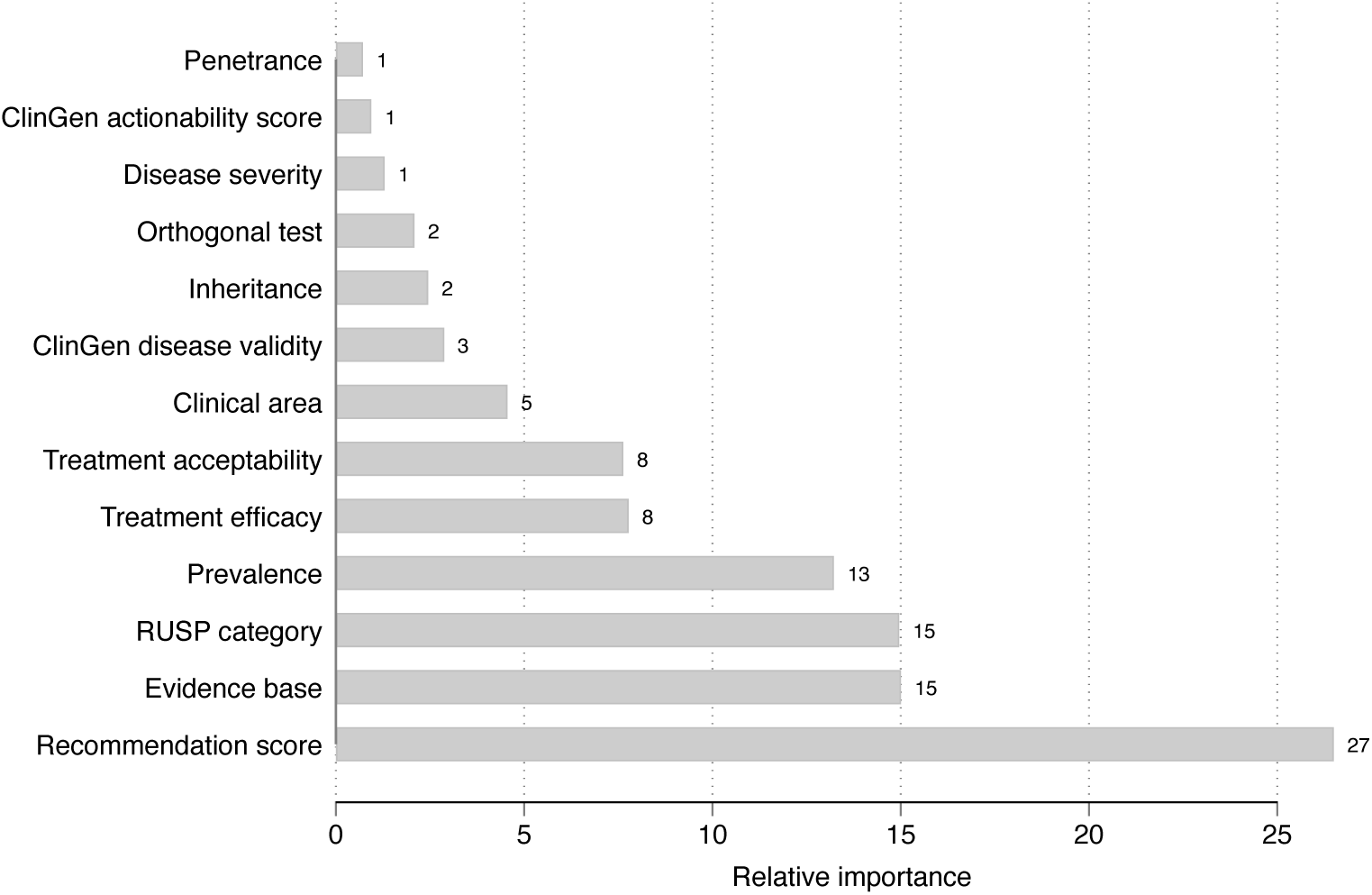
Variable importance in boosted trees model. **Notes:** Bar chart showing the relative importance of variables in the boosted trees model, measured by the reduction in deviance.

**Supplementary Table 1.** Research and commercial genomic newborn screening programs.

|  | Project name | Country | Status (Aug 2024) | Sequencing type | Intended sample size | Unique gene names in list |
| --- | --- | --- | --- | --- | --- | --- |
| Research studies | BabyDetect* | Belgium | Ongoing | Panel | 40,000 | 405 |
|  | BabyScreen+* | Australia | Ongoing | GS/ES | 1,000 | 605 |
|  | BabySeq* | Massachusetts, USA | Ongoing | GS | 100,000 | 4,299 |
|  | BeginNGS* | California, USA | Ongoing | GS/ES | 2,000 | 357 |
|  | Chen et al. 2023* | China | Completed | Panel | 29,601 | 142 |
|  | Cradle | Netherlands | Expected |  |  |  |
|  | Early Check* | North Carolina, USA | Ongoing | GS/ES | 10,000 | 169 |
|  | FirstSteps* | Greece | Ongoing | GS/ES | 101,000 | 385 |
|  | Generation study* | England | Ongoing | GS | 100,000 | 479 |
|  | gnSTAR* | China | Completed | Panel | 4,986 | 134 |
|  | GUARDIAN study* | New York, USA | Ongoing | GS/ES | 100,000 | 314 |
|  | Jian et al. 2022* | China | Completed | GS | 321 | 251 |
|  | Lee et al. 2019* | South Korea | Completed | Panel | 48 | 198 |
|  | Luo et al. 2020* | China | Completed | Panel | 1,127 | 569 |
|  | NeoExome* | China | Completed | Panel | 3,423 | 601 |
|  | NeoSeq* | China | Completed | Panel | 196 | 135 |
|  | NESTS* | China | Completed | Panel | 11,484 | 465 |
|  | New Lives | Germany | Expected |  |  |  |
|  | Newborn Genomics Prog | New Zealand | Expected |  |  |  |
|  | NewbornsInSA* | Australia | Expected | GS | 1,000 | 613 |
|  | Perigenomed | France | Expected | GS | 20,000 |  |
|  | Progetto Genoma Puglia* | Italy | Expected | GS | 6,000 | 404 |
|  | Screen4Care* | European Union | Expected | Panel | 18,000 | 245 |
|  | ScreenPlus | New York, USA | Completed | Panel |  |  |
|  | Sidra Medicine | Qatar | Expected |  |  |  |
|  | TRAIL | Australia | Expected |  |  |  |
|  | Wang et al. 2023* | China | Completed | Panel | 10,224 | 162 |
| Commercial programs | FORESITE 360* | USA | Ongoing | GS |  | 306 |
|  | Fulgent* | USA | Ongoing | Panel |  | 255 |
|  | Igenomix* | Spain | Ongoing | Panel |  | 237 |
|  | KIBs | Kenya | Ongoing | Panel |  |  |
|  | Mendelics* | Brazil | Ongoing | Panel |  | 284 |
|  | Nurture Genomics* | USA | Ongoing | GS |  | 412 |
|  | PerkinElmer* | USA | Ongoing | Panel |  | 268 |
|  | Sema4* | USA | Completed | Panel |  | 167 |

**Notes:** Information on various genomic newborn screening research studies and commercial programs. Gene lists from 27 of these programs were included in the analysis (program names denoted with an asterisk). GS, genome sequencing.

**Supplementary Table 2.** Inclusion criteria employed by various programs for selecting gene-disease associations for genomic newborn screening.

| PROJECT | INCLUSION CRITERIA | SOURCE |
| --- | --- | --- |
| BabyDetect | "General inclusion criteria for genes incorporated into the test panel were the following: significant life-expectancy consequences or severe disability associated with untreated patients, disease onset before 5 years of age, strong genotype-phenotype correlation, existence of a disease-modifying treatment accessible to the diagnosed patients, and significant benefit of prompt treatment. Genes with mutations that underlie diseases currently screened in our biochemical panel were also included even if they did not match these criteria (e.g., G6PD deficiency). Endorsement by pediatricians who specialized in treatment of the disease was mandatory. Screening for a given disease was discontinued if a disease-modifying treatment became unavailable or due to operational issues such as an unacceptable rate of false positives. Discussions were held with experts periodically to review the list of disorders and genes in the tNGS panel." | Boemer et al. 2025 |
| BabyScreen+ | "Genes included in the screening meet the following criteria:<br>A. Screening can be performed with confidence using currently available technology<br>B. The condition associated with the gene has a serious impact on health and/or is life-limiting<br>C. The condition associated with the gene usually affects children under 5 years of age<br>D. There is available treatment that prevents, cures or significantly reduces the severity of the condition" | https://babyscreen.mcri.edu.au/conditions-screened/ |
| BabySeq | The BabySeq study uses a gene filtration and selection approach to generate a list of >6,000 genes disease associations present in ClinVar, HGMD or OMIM, which is then further filtered to approximately ~4,300 genes with stronger association to disease. | Lazo de la Vega et al. 2021 |
| BeginNGS | "Two mandatory criteria<br>1. Severe, childhood-onset disease<br>2. Effective therapeutic intervention available" | Owen et al. 2022 |
| Chen et al. 2023 | "One of the following criteria<br>A. Diseases in the current NBS program in China<br>B. Diseases on the first national list of rare diseases in China<br>C. Diseases on the list of major rare diseases in Shanghai (2016 Edition)<br>D. Diseases in the NBS program recommended by the American College of Medical Genetics and Genomics<br>E. Treatable inherited disorders with a high carrier frequency in Southeast Asia" | Chen et al. 2023 |
| Early Check | "Panel 1 includes gene-condition pairs with high actionability and early age of action (age at intervention initiation), which makes them comparable to core conditions on the Recommended Uniform Screening Panel (RUSP), a set of conditions the Secretary of the U.S. Department of Health and Human Services (HHS) recommends for inclusion in all state NBS programs. All currently recommended RUSP conditions for which sequencing is informative were automatically included on Early Check Panel 1, and other non-RUSP conditions were evaluated for the panel." | Cope et al. 2024 |
| FirstSteps | "The purpose of FirstSteps is the early detection of potentially treatable diseases, so that they can be treated immediately and their complications can be prevented or minimized.<br>The goal of FirstSteps is to ensure prompt and valid treatment for asymptomatic newborns suffering from genetic diseases, in order to prevent or minimize the complications of the disease." | https://www.firststeps-ngs.gr/, translated from Greek |
| Generation study | "Four mandatory criteria<br>1. There is strong evidence that the genetic variant(s) causes the condition and can be reliably detected<br>2. A high proportion of individuals who have the genetic variant(s) would be expected to have symptoms that would have a debilitating impact on quality of life if left undiagnosed<br>3. Early or pre-symptomatic intervention for the condition has been shown to lead to substantially improved outcomes in children, compared to intervention after the onset of symptoms<br>4. Conditions screened for are only those for which the interventions are equitably accessible for all." | https://www.genomicsengland.co.uk/initiatives/newborns/choosing-conditions |
| gnSTAR | "Seventy four inborn disorders were included in this panel based on the Secretary's Advisory Committee on Heritable Disorders in Newborns and Children and the Wilson and Jungner criteria, disorders that can be detected by | Huang et al. 2022 |
|  | biochemical test were selected with consideration of Recommended Uniform Screening Panel (RUSP) recommendation, and disease with high incidences but cannot be screened by C-NBS in China were also enrolled." |  |
| GUARDIAN study | Genes associated with conditions with penetrance >90% before age 5 years and/or for which presymptomatic treatment would be recommended before age 5 years. | Ziegler et al. 2024 |
| Jian et al. 2022 | "Genes associated with 59 Mendelian disorders, 164 primary immunodeficiency diseases (PIDs) and five pharmacogenetic (PGx) genes, following the guidelines by the Recommended Uniform Screening Panel (RUSP), the International Union of Immunologic Societies (IUIS) Expert Committee for Primary Immunodeficiency, the Dutch Pharmacogenetics Working Group (DPWG), and the Clinical Pharmacogenetics Implementation Consortium (CPIC)." | Jian et al. 2022 |
| Lee et al. 2019 | "Actionable child-onset-genes related to NBS was designed for diagnosis of a rare genetic disorder with low incidence before clinical symptoms occurred." | Lee et al. 2019 |
| NeoExome | "To design the NBS NGS panel covering monogenic diseases that are severe, actionable, and early onset for the Chinese population, we combined the following categorization of diseases in our study: 1) 248 related genes (249 diseases) that were included in China National Catalog of Rare Diseases. This catalog includes diseases with low incidence, high risk, and strong treatability based on the clinical data; 2) 360 genes (312 diseases) that were included in Preventable and Treatable Rare Diseases in China; 3) 79 genes (71 diseases) that were now in the conventional MS/MS NBS project in China; 4) 71 genes (71 diseases) that were included in the conventional NBS in China; 5) 149 genes (148 diseases) that were recommended by ACMG; 6) 55 genes (57 diseases) that were recommended by the experts of the Chinese NBS Laboratory Committee. Moreover, a list of diseases with high prevalence in the Chinese population based on the database about positive monogenetic diseases in 40000 ill newborns by MyGenostics was also included. This list included 141 diseases and 94 related genes." |  |
| NESTS | "After curation, we selected 465 causative genes for 596 early-onset, relatively high incidence, and potentially actionable severe inherited diseases in our initiative panel." | Hao et al. 2022 |
| NewbornsInS A | "The 600+ genetic conditions which we will look for meet these criteria:<br>A. Condition occurs in childhood<br>B. The condition is serious, and is expected to have symptoms that would have a debilitating impact on quality of life if undiagnosed<br>C. There is an effective treatment or management option available<br>D. There is strong evidence that the genetic variant causes the condition<br>E. Our laboratory can reliably detect the condition." | https://www.wch.sa.gov.au/research/newbornsinsa-research-study |
| Progetto Genoma Puglia | Genes selected according to age of early onset, significant mortality/morbidity, available treatments and proven effectiveness of early diagnosis in the best/correct care pathway. | ICoNS 2024 |
| Screen4Care | Five inclusion criteria, including treatability, disease onset, disease severity, penetrance, clinical validity. | Internal document |
| FORESITE 360 | The test targets detectable genetic alterations associated with treatable childhood-onset conditions. | https://www.foregenomics.com/products/pediatric-genetic-health-screen |
| Fulgent | Genetic test that identifies DNA changes that can cause infants to develop severe or life-altering conditions. Screen for these conditions, where early detection, intervention, and management are essential for the infant's overall health and quality of life. | https://www.fulgentgenetics.com/products/newborn/newborn.html |
| Igenomix | These genes are responsible for developmental, genetic and metabolic disorders that cause serious health problems starting in early childhood. The ultimate benefit is an early intervention to prevent intellectual and physical disabilities as well as life-threatening illnesses. | https://www.igenomix.eu/genomics-precision-diagnostic/newborn-screening/ |
| Nurture Genomics | Actionable, childhood-onset conditions, considering:<br>• Are most reported cases very early childhood-onset?<br>• Is the natural history of the disease known?<br>• Does early diagnosis result in better outcomes?<br>• Does a treatment or management of the condition exist? | https://nurturegenomics.com/ |
|  | • Is the condition able to be further characterized using non-genetic (orthogonal) testing? |  |
| PerkinElmer | 268 nuclear genes curated for association with medically actionable childhood-onset conditions. | Balciuniene et al. 2023 |
| Sema4 | “Conditions that have onset in infancy or early childhood and for which there is treatment or medical management that, when administered early in an infant or child's life, will significantly improve the clinical outcome. Conditions included in this panel were curated based on criteria such as: inclusion on current state mandated newborn screening panels, onset of symptoms occurring <10 years of age, evidence of high penetrance (>80%), and availability of a treatment or improvement in life due to early intervention.” | Sema4 Natalis Action Sheet |

**Notes:** The criteria enclosed in quotation marks (“”) were directly copied from the specified source material to ensure accuracy and preserve the original wording as presented by the authors.

**Supplementary Table 3.**
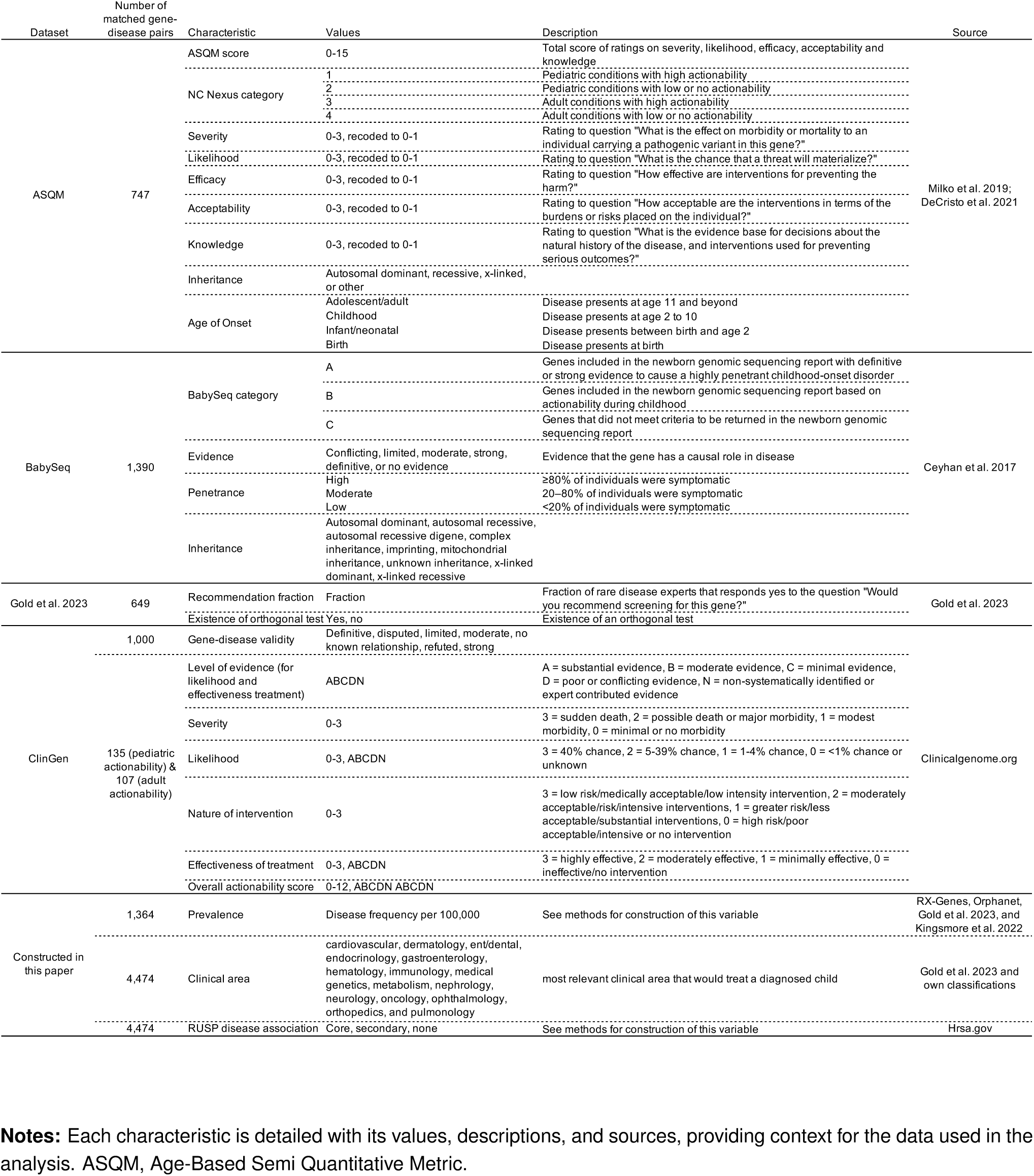
Gene-disease characteristic descriptions.

**Supplementary Table 4.** Gene list summary statistics.

|  | Genes included in<br>at least one gene<br>list | Average<br>across<br>pilots | BabyDetect | BabyScreen+ | BabySeq2 | BeginNGS | Chen et al. | Early<br>Check | FirstSteps | FORESIT<br>E 360 | Fulgent | Generation | gnSTAR | GUARDIAN |
| --- | --- | --- | --- | --- | --- | --- | --- | --- | --- | --- | --- | --- | --- | --- |
| All genes | 4390 | 476 | 405 | 605 | 4299 | 357 | 142 | 169 | 385 | 306 | 255 | 479 | 134 | 314 |
| Clinical area |  |  |  |  |  |  |  |  |  |  |  |  |  |  |
| Cardiovascular | 248 | 22 | 20 | 18 | 243 | 11 | 1 | 4 | 11 | 34 | 8 | 18 | 0 | 6 |
| Dermatology | 104 | 5 | 0 | 2 | 103 | 0 | 0 | 0 | 0 | 1 | 0 | 0 | 0 | 0 |
| ENT/Dental | 146 | 19 | 1 | 54 | 142 | 0 | 0 | 33 | 0 | 15 | 16 | 1 | 2 | 2 |
| Endocrinology | 352 | 53 | 44 | 105 | 348 | 42 | 15 | 22 | 43 | 40 | 38 | 90 | 14 | 29 |
| Gastroenterology | 77 | 10 | 13 | 16 | 75 | 4 | 4 | 4 | 4 | 7 | 6 | 19 | 4 | 6 |
| Hematology | 195 | 34 | 55 | 55 | 183 | 54 | 8 | 8 | 58 | 13 | 8 | 44 | 3 | 15 |
| Immunology | 274 | 57 | 64 | 122 | 250 | 69 | 12 | 10 | 70 | 26 | 26 | 106 | 11 | 96 |
| Medical Genetics | 556 | 32 | 7 | 21 | 546 | 7 | 1 | 5 | 7 | 12 | 11 | 9 | 4 | 1 |
| Metabolism | 492 | 121 | 129 | 138 | 480 | 108 | 87 | 67 | 119 | 99 | 98 | 89 | 90 | 119 |
| Nephrology | 122 | 15 | 9 | 17 | 120 | 11 | 4 | 7 | 14 | 10 | 10 | 17 | 0 | 11 |
| Neurology | 1251 | 74 | 51 | 36 | 1242 | 47 | 8 | 5 | 54 | 20 | 18 | 37 | 3 | 23 |
| Oncology | 126 | 10 | 2 | 16 | 122 | 1 | 1 | 2 | 1 | 23 | 11 | 14 | 2 | 2 |
| Ophthalmology | 259 | 13 | 8 | 2 | 258 | 0 | 0 | 0 | 1 | 4 | 3 | 1 | 0 | 2 |
| Orthopedics | 119 | 6 | 0 | 2 | 119 | 1 | 0 | 0 | 1 | 1 | 1 | 0 | 1 | 1 |
| Pulmonology | 67 | 6 | 2 | 1 | 66 | 2 | 1 | 2 | 2 | 1 | 1 | 34 | 0 | 1 |
| RUSP |  |  |  |  |  |  |  |  |  |  |  |  |  |  |
| Core | 48 | 41 | 46 | 45 | 48 | 43 | 35 | 40 | 42 | 44 | 44 | 46 | 38 | 46 |
| Secondary | 39 | 27 | 33 | 24 | 38 | 28 | 21 | 14 | 27 | 27 | 25 | 18 | 28 | 30 |
| ASQM |  |  |  |  |  |  |  |  |  |  |  |  |  |  |
| Genes | 741 |  | 240 | 340 | 732 | 194 | 114 | 168 | 211 | 269 | 225 | 255 | 112 | 194 |
| Average ASQM score | 10.30 |  | 11.15 | 11.52 | 10.32 | 11.49 | 11.57 | 12.42 | 11.41 | 11.62 | 11.68 | 11.50 | 11.62 | 11.30 |
| BabySeq |  |  |  |  |  |  |  |  |  |  |  |  |  |  |
| Genes | 1337 |  | 253 | 365 | 1328 | 230 | 130 | 151 | 253 | 263 | 221 | 281 | 116 | 205 |
| Genes in 3 categories |  |  |  |  |  |  |  |  |  |  |  |  |  |  |
| A | 826 |  | 204 | 300 | 826 | 172 | 120 | 135 | 192 | 217 | 204 | 228 | 104 | 179 |
| B | 59 |  | 11 | 8 | 59 | 11 | 3 | 7 | 11 | 29 | 5 | 5 | 5 | 7 |
| C | 452 |  | 38 | 57 | 443 | 47 | 7 | 9 | 50 | 17 | 12 | 48 | 7 | 19 |
| Gold et al. 2023 |  |  |  |  |  |  |  |  |  |  |  |  |  |  |
| Genes | 644 |  | 257 | 388 | 622 | 274 | 74 | 65 | 305 | 139 | 122 | 317 | 50 | 214 |
| Avg recommendation | 56.24 |  | 63 | 61 | 56 | 60 | 71 | 73 | 61 | 67 | 68 | 60 | 71 | 63 |

|  | Igenomix | Jian et al. | Lee et al. | Luo et al. | Mendelics | NeoExome | NeoSeq | NESTS | NewbornsInSA | Nurture | PerkinElmer | Prog. Puglia | Screen4Care | Sema4 | Wang et al. |
| --- | --- | --- | --- | --- | --- | --- | --- | --- | --- | --- | --- | --- | --- | --- | --- |
| All genes | 237 | 251 | 198 | 569 | 284 | 601 | 135 | 465 | 613 | 412 | 268 | 404 | 245 | 167 | 162 |
| Clinical area |  |  |  |  |  |  |  |  |  |  |  |  |  |  |  |
| Cardiovascular | 2 | 11 | 12 | 31 | 7 | 50 | 0 | 23 | 21 | 14 | 18 | 11 | 4 | 3 | 1 |
| Dermatology | 0 | 1 | 0 | 8 | 0 | 7 | 0 | 6 | 2 | 0 | 0 | 0 | 0 | 0 | 0 |
| ENT/Dental | 12 | 8 | 3 | 41 | 2 | 6 | 2 | 84 | 54 | 1 | 15 | 3 | 0 | 0 | 10 |
| Endocrinology | 37 | 8 | 23 | 41 | 47 | 73 | 14 | 23 | 106 | 53 | 38 | 52 | 33 | 27 | 14 |
| Gastroenterology | 6 | 2 | 3 | 10 | 17 | 17 | 4 | 5 | 16 | 10 | 7 | 6 | 1 | 6 | 4 |
| Hematology | 6 | 6 | 5 | 54 | 15 | 58 | 3 | 28 | 56 | 65 | 7 | 54 | 36 | 8 | 8 |
| Immunology | 24 | 118 | 4 | 34 | 52 | 40 | 11 | 30 | 122 | 77 | 26 | 73 | 28 | 13 | 12 |
| Medical Genetics | 1 | 17 | 21 | 58 | 0 | 39 | 4 | 37 | 21 | 6 | 13 | 5 | 2 | 1 | 3 |
| Metabolism | 126 | 66 | 81 | 142 | 107 | 169 | 91 | 87 | 139 | 135 | 99 | 138 | 96 | 86 | 90 |
| Nephrology | 6 | 0 | 7 | 15 | 11 | 25 | 0 | 34 | 18 | 17 | 10 | 10 | 6 | 8 | 4 |
| Neurology | 14 | 6 | 23 | 96 | 23 | 83 | 3 | 40 | 36 | 22 | 16 | 45 | 33 | 12 | 13 |
| Oncology | 0 | 5 | 2 | 9 | 1 | 12 | 2 | 4 | 16 | 9 | 14 | 5 | 2 | 2 | 1 |
| Ophthalmology | 1 | 1 | 9 | 15 | 1 | 14 | 0 | 32 | 3 | 1 | 3 | 1 | 1 | 0 | 0 |
| Orthopedics | 1 | 1 | 2 | 9 | 0 | 5 | 1 | 4 | 2 | 0 | 1 | 0 | 2 | 0 | 1 |
| Pulmonology | 1 | 1 | 3 | 6 | 1 | 3 | 0 | 28 | 1 | 2 | 1 | 1 | 1 | 1 | 1 |
| RUSP |  |  |  |  |  |  |  |  |  |  |  |  |  |  |  |
| Core | 43 | 45 | 27 | 44 | 33 | 44 | 38 | 33 | 45 | 39 | 41 | 44 | 39 | 44 | 35 |
| Secondary | 34 | 34 | 20 | 33 | 21 | 38 | 29 | 29 | 24 | 20 | 25 | 37 | 25 | 27 | 21 |
| ASQM |  |  |  |  |  |  |  |  |  |  |  |  |  |  |  |
| Genes | 196 | 136 | 126 | 316 | 168 | 301 | 113 | 277 | 346 | 235 | 236 | 242 | 176 | 167 | 125 |
| Average ASQM score | 11.26 | 11.12 | 11.23 | 10.96 | 11.54 | 10.82 | 11.59 | 11.09 | 11.51 | 11.23 | 11.67 | 11.32 | 11.94 | 11.69 | 11.66 |
| BabySeq |  |  |  |  |  |  |  |  |  |  |  |  |  |  |  |
| Genes | 190 | 129 | 168 | 462 | 177 | 415 | 117 | 364 | 372 | 256 | 233 | 268 | 216 | 142 | 142 |
| Genes in 3 categories |  |  |  |  |  |  |  |  |  |  |  |  |  |  |  |
| A | 166 | 106 | 139 | 385 | 155 | 312 | 104 | 278 | 304 | 209 | 204 | 215 | 189 | 129 | 130 |
| B | 5 | 5 | 10 | 19 | 3 | 29 | 5 | 15 | 10 | 10 | 16 | 10 | 6 | 4 | 3 |
| C | 19 | 18 | 19 | 58 | 19 | 74 | 8 | 71 | 58 | 37 | 13 | 43 | 21 | 9 | 9 |
| Gold et al. 2023 |  |  |  |  |  |  |  |  |  |  |  |  |  |  |  |
| Genes | 111 | 88 | 57 | 169 | 183 | 233 | 50 | 119 | 393 | 347 | 125 | 313 | 161 | 85 | 79 |
| Avg recommendation | 70 | 57 | 67 | 65 | 65 | 61 | 71 | 62 | 61 | 65 | 68 | 64 | 67 | 70 | 70 |

**Notes:** The upper part of the table shows the number of genes by clinical area. The ASQM, BabySeq, and Gold et al. 2023 sections display the number of genes each program has in common with these datasets, along with summary statistics. Averages are calculated as means. The RUSP category indicates whether genes are in the Core or Secondary panel. ASQM, Age-Based Semi Quantitative Metric; RUSP, Recommended Uniform Screening Panel.

**Supplementary Table 5.** Univariate gene list regressions results.

|  | (1) | (2) | (3) | (4) | (5) | (6) | (7) | (8) | (9) |
| --- | --- | --- | --- | --- | --- | --- | --- | --- | --- |
| Dependent variable: gene list inclusion fraction |  |  |  |  |  |  |  |  |  |
| RUSP (base: not in RUSP) |  |  |  |  |  |  |  |  |  |
| Core | 0.747***<br>(0.019) | 0.521***<br>(0.032) | 0.576***<br>(0.034) | 0.608***<br>(0.033) | 0.590***<br>(0.033) | 0.606***<br>(0.033) | 0.546***<br>(0.031) | 0.330***<br>(0.065) | 0.292***<br>(0.058) |
| Secondary | 0.600***<br>(0.022) | 0.367***<br>(0.035) | 0.452***<br>(0.037) | 0.461***<br>(0.037) | 0.464***<br>(0.036) | 0.463***<br>(0.037) | 0.451***<br>(0.034) | 0.271***<br>(0.102) | 0.320***<br>(0.092) |
| Gene-disease characteristics |  |  |  |  |  |  |  |  |  |
| Neonatal or infant onset |  | 0.152***<br>(0.023) |  |  |  |  |  |  |  |
| Severity |  |  | 0.145***<br>(0.032) |  |  |  |  |  |  |
| Penetrance |  |  |  | 0.155***<br>(0.029) |  |  |  |  |  |
| Efficacy |  |  |  |  | 0.170***<br>(0.024) |  |  |  |  |
| Acceptability |  |  |  |  |  | 0.150***<br>(0.023) |  |  |  |
| Evidence base |  |  |  |  |  |  | 0.295***<br>(0.025) |  |  |
| Existence of an orthogonal test |  |  |  |  |  |  |  | 0.138***<br>(0.023) |  |
| Expert recommendations<br>≥80% Yes |  |  |  |  |  |  |  |  | 0.435***<br>(0.031) |
| Constant | 0.093***<br>(0.002) | 0.209***<br>(0.019) | 0.168***<br>(0.019) | 0.109***<br>(0.026) | 0.133***<br>(0.018) | 0.133***<br>(0.020) | 0.077***<br>(0.016) | 0.184***<br>(0.021) | 0.273***<br>(0.007) |
| Observations | 4,475 | 747 | 749 | 749 | 749 | 749 | 749 | 637 | 649 |
| R-squared | 0.333 | 0.473 | 0.394 | 0.401 | 0.418 | 0.410 | 0.478 | 0.105 | 0.278 |
| Standard errors in parentheses |  |  |  |  |  |  |  |  |  |
| *** p<0.01, ** p<0.05, * p<0.1 |  |  |  |  |  |  |  |  |  |

**Notes:** Coefficients and standard errors (in parentheses) from univariate linear regressions, with the dependent variable being the gene list inclusion fraction (proportion of programs including each gene, between 0 and 1).

**Supplementary Table 6.**
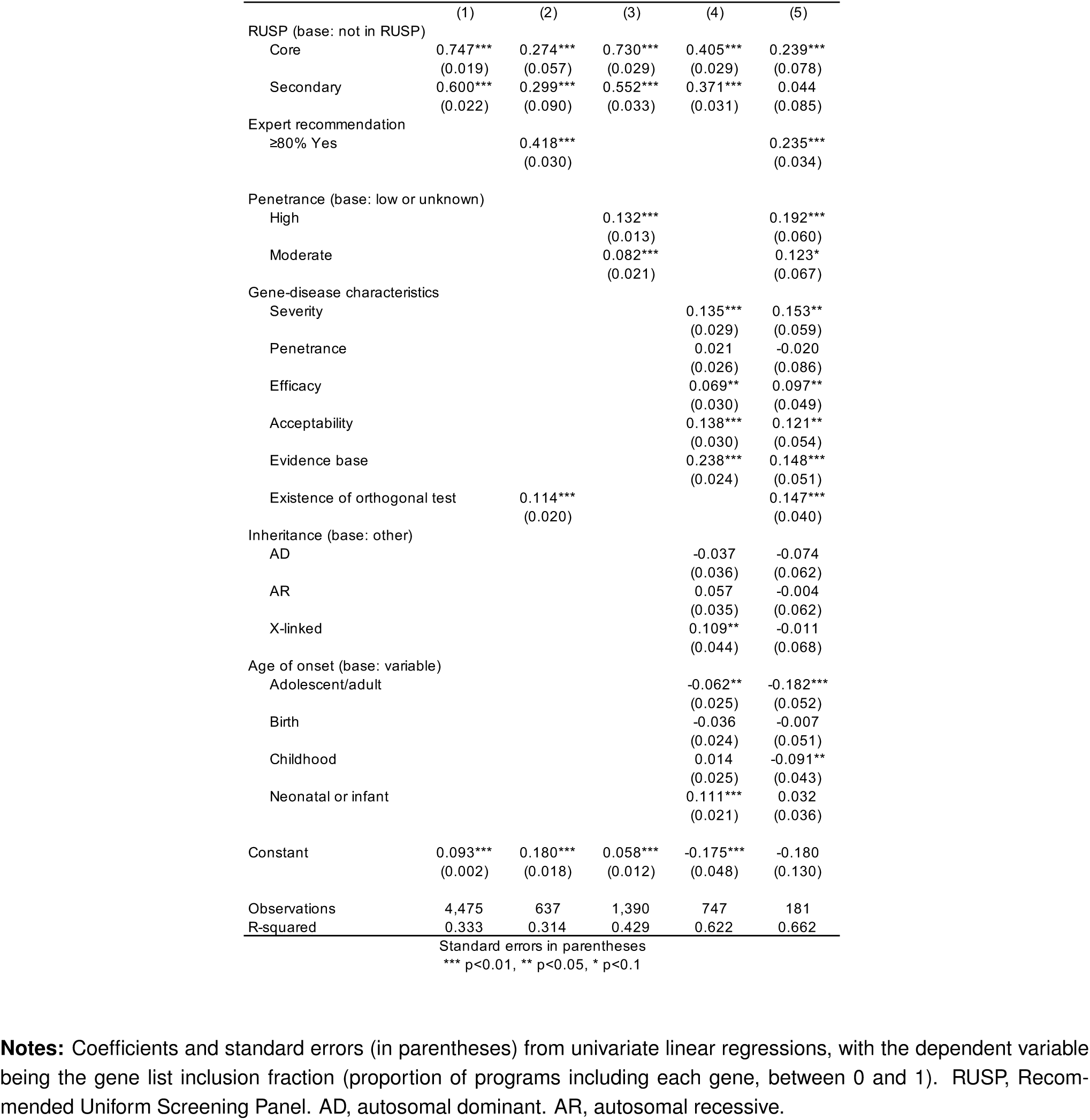
Multivariate gene list inclusion regressions.

**Supplementary Table 7.**
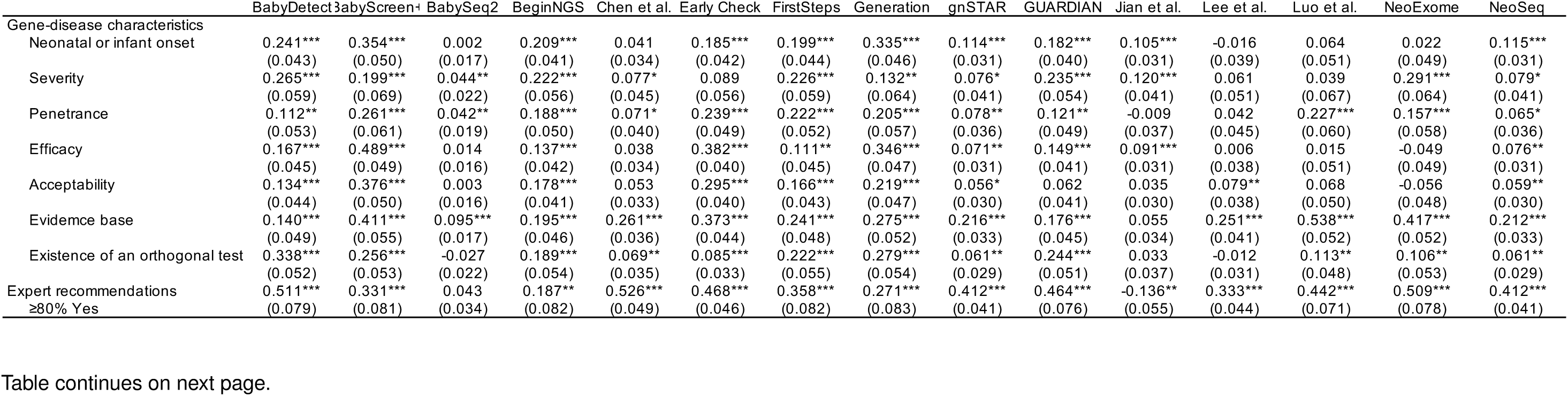
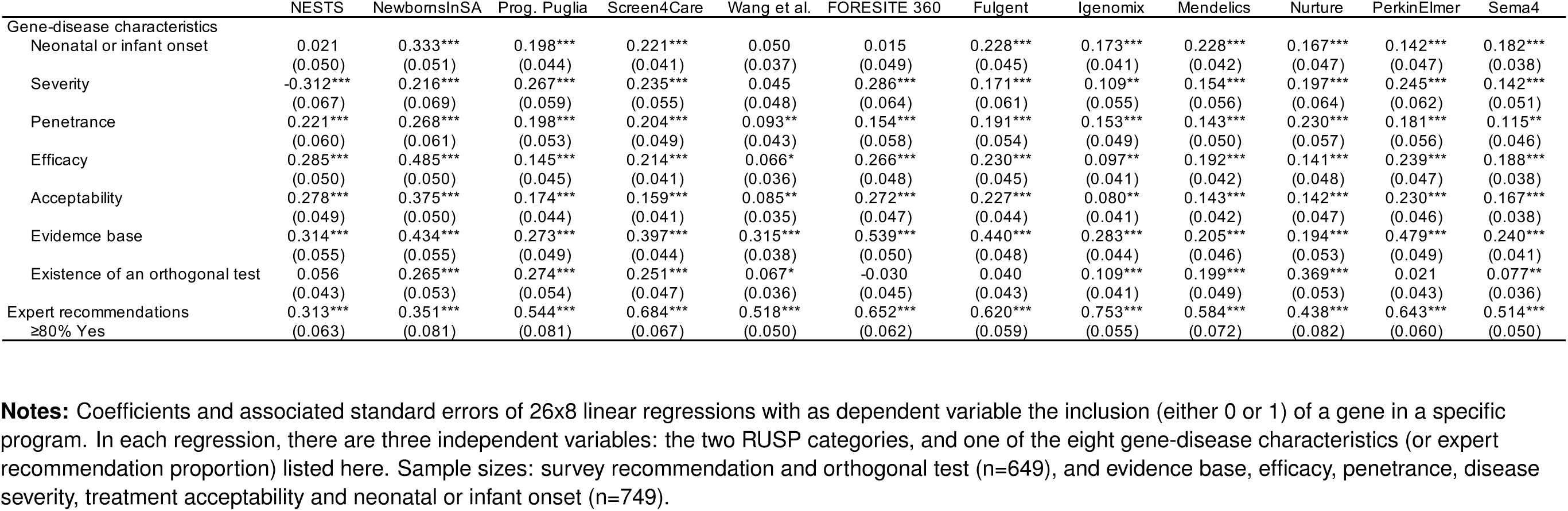
Univariate gene list regression results by program.

## Notes

### Summary of Updates

Updated and revised manuscript text. Table 1 updated with new information. Tabel 2 updated after re-running algorithm with updated gene list. Updated Figures 1,2,3 after updating gene list.

